# Attenuation of typical sex differences in the time-resolved functional connectivity of the fusiform gyrus in autism

**DOI:** 10.64898/2026.06.02.26354318

**Authors:** Dorothea L. Floris, Luigi F. Saccaro, Farnaz Delavari, Dawid Strzelczyk, Bruno Hebling Vieira, Camille Elleaume, Charlotte M. Pretzsch, Christine Ecker, Tobias Banaschewski, Rosemary J. Holt, Simon Baron-Cohen, Thomas Bourgeron, Tony Charman, Eva Loth, Declan G. M. Murphy, Jan K. Buitelaar, Christian F. Beckmann, Dimitri Van De Ville, APEX Consortium, EU-AIMS LEAP group, Nicolas Langer

## Abstract

**Background:** Autism is characterized by social-communicative difficulties, with sex differences in symptom presentation. Social functioning is inherently dynamic, however, many neuroimaging studies rely on static, time-averaged approaches that obscure time-varying network interactions, potentially limiting our ability to capture the dynamic processes underlying social cognition. The fusiform gyrus (FFG), central to face and social perception, shows differences in functional connectivity in autism, yet is rarely examined dynamically or as a spatially heterogeneous structure. Here, we investigate the dynamic functional connectivity of FFG subregions in terms of their large-scale network configurations as a function of diagnosis and sex.

**Methods:** We applied micro co-activation patterns analysis (μCAPs) to resting-state fMRI data from 286 autistic individuals (208:78 males:females) and 228 non-autistic individuals (146:82 males:females), aged 6-30 years, from the EU-AIMS LEAP dataset. μCAPs were identified using k-means clustering with FFG as the seed, and connectopic mapping positioned each μCAP along the principal connectivity gradient. We quantified μCAPs occurrence and further examined dwell time, transition probabilities, and spatial extent, along with associations with social functioning.

**Results:** Six μCAPs mapped onto distinct FFG subregions along a posterior-anterior axis. A significant sex-by-diagnosis interaction emerged for a default mode network (DMN)-related μCAP. Non-autistic females exhibited significantly more frequent occurrences, longer dwell times and distinct transition dynamics compared to males, while no sex difference was observed in autism. The spatial extent of this μCAP showed a reversal of typical sex effects.

**Conclusions:** Autism is associated with an attenuation and reversal of typical sex differences in the functional configuration and spatial extent of FFG-DMN coupling, indicating that neural signatures of social-cognitive functions are sex-specific and dynamic. These findings suggest that sex is a neurobiologically meaningful dimension of heterogeneity in autism, expressed in dynamic network organization.

## Introduction

Autism spectrum disorder (‘autism’) is a neurodevelopmental condition characterized by difficulties in social communication and restricted, repetitive behaviours. Challenges in social functioning are among the most consequential features, impacting quality of life, academic performance and social integration throughout the lifespan [1, 2]. Although social-cognitive interventions can yield benefits, their efficacy is often modest and variable [3, 4], in part because they are not optimally tailored to individuals’ underlying neurobiological profiles. Crucially, social functioning in everyday life is inherently dynamic, requiring individuals to continuously adjust and adapt to changing social contexts, yet much of the existing neuroimaging literature has characterized the neural substrates of social processing using static, time-averaged approaches that obscure how functional interactions dynamically reconfigure over time. Thus, a better understanding of the dynamic neurobiological mechanisms contributing to variability in social functioning is of utmost importance for informing more targeted and effective interventions.

Growing evidence indicates that autism presents differently in males and females, with diverging symptom profiles [5, 6], developmental trajectories [7], age of diagnosis [8] and polygenic architecture [9]. These sex differences contribute substantially to the broader heterogeneity of the autism spectrum. Despite growing neuroimaging evidence of sex-related differences in brain structure [10, 11] and function [12, 13], however, their neural basis remains poorly understood. Given that autism has been conceptualized as involving widespread differences in brain connectivity [14, 15], resting-state functional magnetic resonance imaging (rs-fMRI) has been widely used to study sex-related variation in large-scale functional networks [12, 13, 16–22]. These studies consistently implicate the default mode (DMN), salience, sensorimotor, and attention networks, though reported patterns vary, aligning in some cases with sex-atypical profiles, in others with a shift toward neurotypical male-like organization [12, 13, 16, 17, 20–24]. Despite this variability, many findings converge on circuitry supporting social cognition. Particularly autistic females have been shown to exhibit reduced functional connectivity (FC) in the DMN [23, 25] which is central to social cognition and mentalizing, along with sex-related connectivity patterns linked to both camouflaging behaviours [26] and reward-prefrontal circuits associated with social functioning [27]. Additionally, FC within face-processing regions shows attenuated sex differences in autistic children [18]. These findings suggest that sex differences in autism may be particularly pronounced in brain systems underlying social information processing. Elucidating their neurobiological bases could ultimately inform more tailored approaches to intervention and support.

Among social-cognitive processes, face processing has been frequently implicated as a domain where autistic individuals may experience difficulties [28, 29]. Research into the neural bases of face processing has identified a distributed bilateral neural network of specialized brain regions. Here, the fusiform gyrus (FFG) is the largest structure in the higher-level visual cortex, characterized by marked functional heterogeneity [30] and selective responses to facial stimuli [31, 32]. In autism, the FFG has frequently been implicated with studies showing differences in FC [33–35] and structural properties, including volume [36] and asymmetry [37], as well as altered electrophysiological [38, 39] and fMRI task-evoked responses [40–42]. We recently also reported a multimodal neural signature within the FFG that differed on average between autistic and non-autistic individuals and was associated with individual differences in social-cognitive functioning within the autistic group [43].

The FFG exhibits a functional gradient along its anterior–posterior axis, with posterior medial regions primarily connected to visual and auditory networks, lateral regions to language systems, and anterior regions to the DMN [44]. This organization supports a continuum from lower-level sensory processing to higher-order, domain-specific functions such as face perception [45, 46]. While this functional heterogeneity motivates detailed regional analyses, the FFG is also embedded within distributed brain networks through extensive anatomical and functional connections supporting diverse cognitive functions [44, 45, 47]. Considering this broader connectivity profile may therefore provide a more comprehensive and biologically meaningful account. Consistent with this, in autism, differences in FC within social brain networks, including FFG connectivity, have been linked to increased social difficulties [33, 34], further underscoring the need to examine FFG function within its broader network context.

Such heterogeneous and distributed organization is inherently dynamic, yet most previous FC studies in autism rely on static and time-averaged approaches, which overlook temporal fluctuations and fine-grained spatial heterogeneity, obscuring state-dependent and region-specific effects potentially relevant for sex differences. Dynamic functional connectivity (dFC) methods, most commonly using sliding windows (SW), have increasingly been applied to address these limitations, revealing differences in temporal network organization in autism, including altered variability in DMN [48] and other networks [49–52], and prolonged state dwell times with fewer transitions between connectivity states [53–55]. These dynamic features have been linked to differences in social-cognitive functioning [48, 50, 55, 56]. However, SW approaches remain limited in temporal precision requiring arbitrary selection of window length and having inherent trade-offs between sensitivity and stability [57, 58]. This motivates alternative data-driven frameworks such as coactivation patterns (CAPs) analysis [59, 60], which identifies recurrent brain states directly from selected time frames, yielding temporally precise and more interpretable representations of transient network states while also avoiding reliance on predefined temporal parameters. In recent years, CAPs studies have provided converging evidence that autism is associated with differences in the dynamic engagement of large-scale brain networks. Across whole-brain and network-focused CAPs analyses, autism has been characterized primarily by differences in state frequency and spatial configuration [61–63]. Other CAPs studies in autism report differences in engagement of DMN and salience-related states, reduced engagement of executive and attentional control states, and greater spatial and temporal variability in these transient brain states [64–66], with several studies linking these features to social and clinical symptom severity [61, 64, 65]. Together, these findings underscore the value of CAP-based approaches for capturing clinically meaningful brain dynamics beyond static connectivity. However, whether such dynamic properties differ by sex within functionally distinct subregions crucial to face processing in autism remains to be established.

Notably, these previous CAPs studies have adopted whole-brain perspectives, leaving open the question of how dynamic state properties are expressed within functionally specialized regions related to social perception, such as the FFG. However, given the pronounced functional heterogeneity of the FFG and its widespread interactions with distributed brain networks, treating the FFG as a homogeneous seed to establish CAPs may obscure region-and state-specific differences in autism. Hence, to address this, we employed a recent extension, so-called micro co-activation patterns (μCAPs) analysis [67–69], which, unlike conventional CAPs, allows capturing functional subdivisions within the seed region.

To capture intrinsic, time-varying network organization independent of task demands, we focused on rs-fMRI. Using this approach, we examined sex differences in the dFC of the FFG in autistic and non-autistic individuals. We first identified FFG-related μCAPs and their corresponding functional subdivisions within the FFG, and then characterised these patterns across spatial and temporal domains. Specifically, we assessed whether μCAPs differed in their frequency of dynamic coupling with the FFG across autistic and non-autistic males and females, as well as their finer-grained temporal properties, spatial extent, and associations with social functioning. By moving beyond static, sex-agnostic models of FC, we aim to reveal how sex and autism interact to shape the dynamic neural architecture that underpins social-cognitive heterogeneity in autism.

## Methods and Materials

### Sample Characterisation

Participants were part of the EU-AIMS/AIMS-2-TRIALS Longitudinal European Autism Project (LEAP) cohort [70, 71]. They underwent detailed clinical, cognitive, and MRI assessments at one of six collaborating sites. All autistic participants had an established clinical diagnosis of autism, which was confirmed using information from the gold-standard diagnostic instruments, Autism Diagnostic Interview-Revised [72] (ADI-R) and the Autism Diagnostic Observation Schedule [73] (ADOS). The study was approved by local research ethics committees at each site (via the Integrated Research Application System [IRAS], UK; see Table S1). Written informed consent was obtained from all participants, or from a parent/legal guardian for minors or those unable to provide consent. Participants were compensated for their study visits. Exclusion criteria comprised MRI contraindications, lack of written consent for MRI, and significant hearing or visual impairments not corrected by glasses or hearing aids. Individuals with an IQ below 50 were excluded. In the autistic group, co-occurring psychiatric conditions were permitted except for psychosis or bipolar disorder, whereas in the non-autistic group, self- or parent-reported psychiatric disorders were exclusionary. A history of alcohol or substance abuse/dependence in the past year was an exclusion criterion for both groups. Participants were included if medication use was stable for at least eight weeks prior to and during the baseline assessment. For further details on diagnostic procedure, study design and exclusion criteria, see Supplement and our prior works [70, 71]. For a detailed flowchart depicting the final sample selection, see Figure S1.

### MRI data acquisition and preprocessing

MRI data were acquired at five European sites on 3T scanners using harmonized protocols (see Methods in Supplement, Tables S2 and S3). High-resolution T1-weighted scans and 8–10 min multi-echo rs-fMRI runs (TR = 2.3 s) were collected while participants fixated on a cross. Preprocessing followed standard pipelines (FSL v5.0.6), including motion correction, ICA-AROMA denoising, nuisance regression (white matter/CSF), temporal filtering (0.01 Hz), and spatial normalisation to 2 mm MNI152 space (see Supplement).

### Micro co-activation pattern analysis

Analyses were conducted on rs-fMRI data to examine intrinsic network dynamics. We applied μCAPs analysis, a method that allows for spatial heterogeneity within the seed region. μCAPs builds on the established CAPs framework, which has been widely used and described in prior work [59, 60, 74] and is further detailed in the Supplement. While traditional CAPs analyses assume a uniform seed region, μCAPs iteratively refine the seed, thus allowing differentiation within it for each identified brain state. This enables a more fine-grained characterisation by revealing how its substructures contribute to dynamic brain states. Here, the FFG was selected as the seed region due to its central involvement in social cognition, its established relevance in autism, and its functionally heterogeneous and distributed connectivity profile, which makes it particularly well suited for capturing dynamic connectivity patterns. The ROI was defined from the Harvard-Oxford atlas (HOA) (fMRIB, Oxford, UK) by combining the anterior and posterior temporal fusiform, temporal–occipital fusiform, and occipital fusiform regions bilaterally. The μCAPs method yields k μCAPs, each linked to a spatial seed weight map. Unlike independent component analysis, which decomposes continuous time series into spatially independent components, the μCAPs method identifies recurring patterns of whole-brain co-activation from selected time frames capturing transient and state-dependent network configurations. The optimal number of k was determined using an established cross-validation procedure as in prior work [67]. The μCAPs analysis code is openly available at https://github.com/MIPLabCH/mCAP and is described in detail in the Supplement and elsewhere [67, 68]. For an overview, see Figure 1.

**Figure 1.**
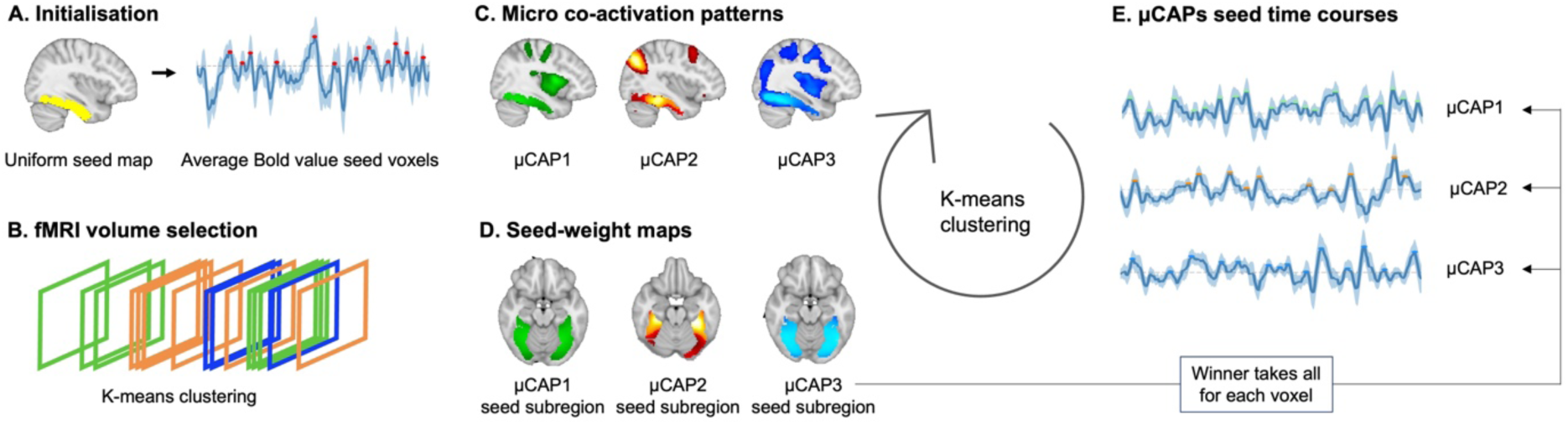
Flowchart of the μCAPs Processing Pipeline. (**A**) Initialisation. The analysis begins with a uniform seed region (top left, shown in yellow). A Z-scored time course of seed activity is computed and in order to identify time points of significant activation it is thresholded for the first iteration at 0.5 to maximize sensitivity. (**B-C**) FMRI volume selection and clustering into μCAPs. The corresponding fMRI volumes are clustered into μCAPs using K-means clustering. (**D**) Seed-weight maps. For each μCAP, a spatial weight map within the seed region is extracted, and voxels are updated by retaining only their maximal activation value across patterns (i.e., winner-takes-all approach). (**E**) Extraction of μCAPs seed time courses. New seed time courses are then computed and for subsequent iterations they are thresholded at 1.0 (a value within the range of typical choices for conventional CAP analyses [59, 75]) to extract a refined set of volumes for the next iteration. This process is repeated until convergence is reached.

### Functional Annotation of μCAPs

To annotate each identified μCAP with a corresponding functional network, we computed its spatial overlap with the 17 large-scale canonical networks defined by Yeo et al. [76] (see Figure 2G). Specifically, each μCAP was overlaid with the Yeo network maps in MNI space, and the μCAP was assigned to the network with which it showed the greatest voxel-wise overlap. To further characterize the identified subregions of the FFG seed, we used the probabilistic functional atlas of human occipito-temporal visual cortex (VIS atlas) [30] (Figure 3C). The atlas was intersected with the FFG seed mask, so that we obtained VIS atlas subregions corresponding to early visual and category-selective areas (e.g., faces, bodies, characters, places). This enabled a regionally specific mapping of seed contributions, identifying which functional subareas within the FFG were differentially involved across the identified μCAPs.

**Figure 2.**
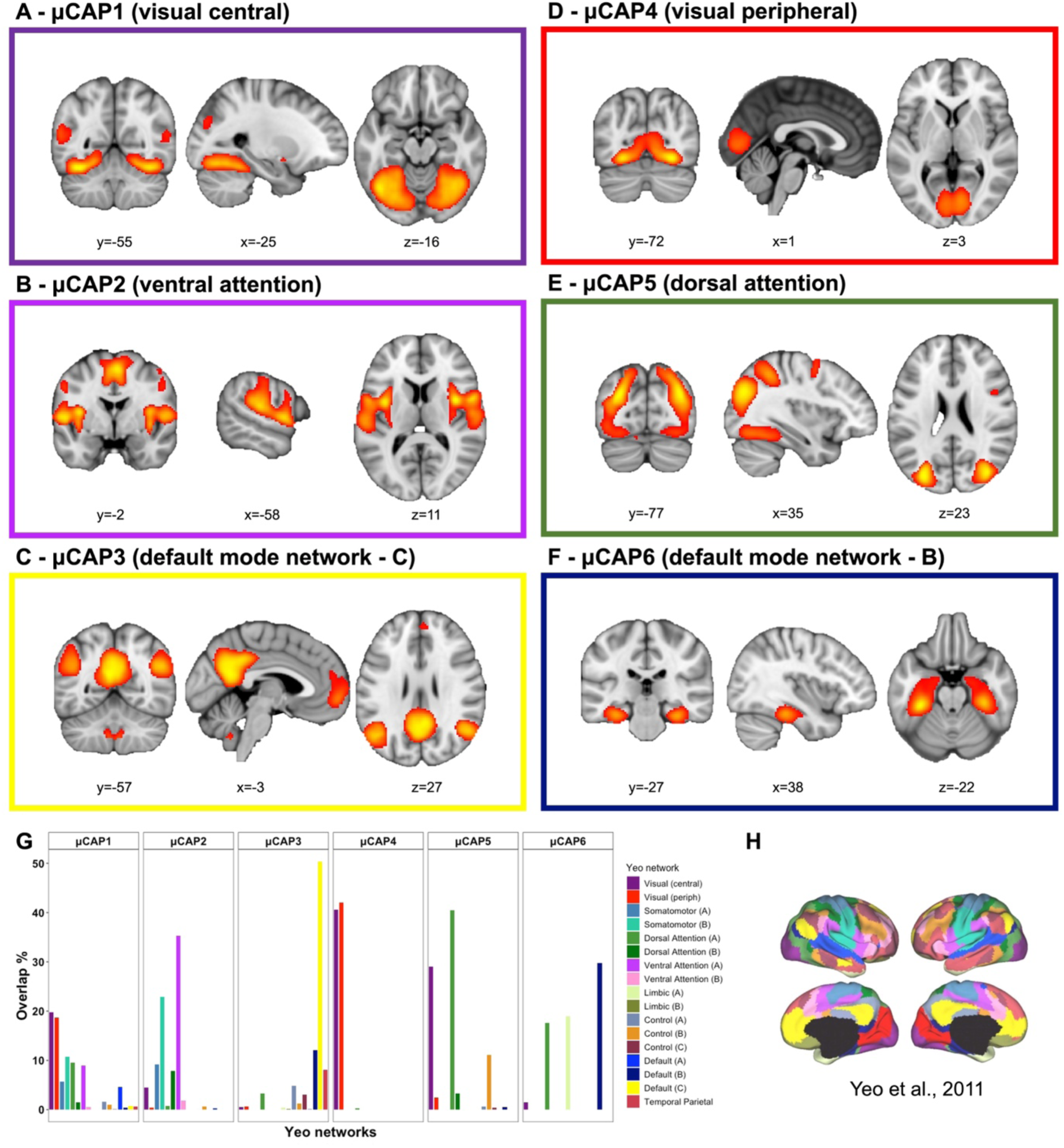
μCAP Networks. Six μCAPs identified using the fusiform gyrus (FFG) as the seed region. Panels **A–F** show the spatial maps of the six μCAPs (thresholded at the 90th percentile): a visual-central μCAP (**A**), a ventral attention μCAP (**B**), a default mode network (DMN) - C μCAP (**C**), a visual-peripheral μCAP (**D**), a dorsal attention μCAP (**E**) and a DMN - B μCAP (**F**). Functional network annotations were obtained by computing the percentage of spatial overlap between each μCAP and the 17 canonical Yeo networks (**H**). Panel **G** displays the 17 functional networks and subdivisions (e.g., A, B, C) from Yeo et al. (2011), used as the reference for network labeling.

**Figure 3.**
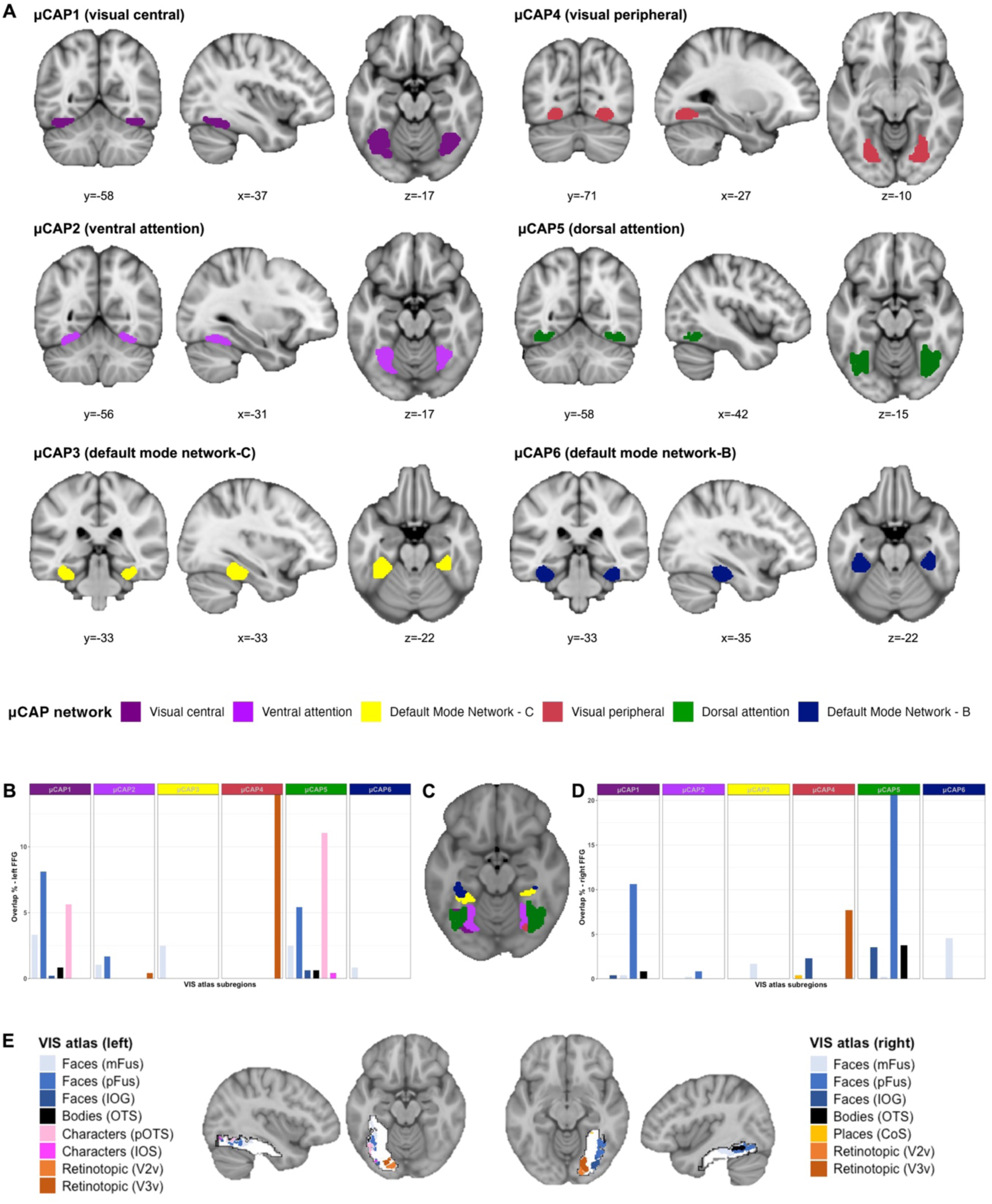
FFG Subregions Co-activating with μCAP Networks. Data-driven μCAP sub-regions of the FFG. Panel (**A**) and (**C**) show the six subregions within the FFG that co-activate with the six μCAPs networks color-coded by the Yeo networks. Panel (**B**) and (**D**) show the spatial overlap of suprathreshold voxels (above the 90th percentile) with the probabilistic functional atlas of the occipito-temporal cortex (i.e., VIS atlas [30]), which covers different early visual and category-selective patches within the left and right FFG. Panel (**E**) depicts the VIS atlas and its different category-selective subregions in the left and right hemispheres.

In addition, we computed functional connection topographies within the right and left FFG separately for each individual using connectopic mapping [77] to characterize the intrinsic functional organization of the FFG along a continuous gradient and to assess how μCAP-derived within-seed weight maps are organized along this axis (see Methods in Supplement). This approach complements the discrete μCAP parcellation by situating these subregions within a continuous functional hierarchy, and enabled us to test whether group differences reflect shifts or dispersion along this organizational axis, while also providing an independent validation of μCAP–defined subregions. The resulting maps capture systematic spatial variation in connectivity between the FFG and the rest of the cortex. To characterize each FFG μCAP along the principal functional gradient of intrinsic organization, we derived group-average gradient maps for each hemisphere and divided them into ten ordinal bins based on deciles of the gradient value distributions, spanning the continuum from lower-level visual to higher-order associative regions. Spatial overlap between each gradient bin and the six FFG μCAP parcels was quantified as the percentage of parcel voxels falling within each bin, yielding parcel-specific enrichment profiles across the gradient.

To assess whether μCAP distributions along the FFG gradient varied by sex and diagnosis, the same procedure was applied at the individual level using subject-specific μCAP parcels. Resulting enrichment profiles were analyzed using linear mixed-effects models separately for each μCAP, with gradient bin, diagnosis, and sex as fixed effects, and age, mean framewise displacement (meanFD), and site as covariates, and subject included as a random intercept. P-values for fixed effects of interest were corrected across μCAPs using the Benjamini–Hochberg False Discovery Rate (FDR) procedure. Bin-specific simple effects were assessed using model-derived contrasts with FDR correction across bins.

### Temporal, Clinical and Spatial Characterisation of μCAPs

To temporally characterize the identified μCAPs, we first computed the number of times each μCAP was expressed per participant across the time course of the recording (i.e., occurrences). Occurrence frequency was used as the primary selection metric as it captures the overall prevalence of a given μCAP, which can be further decomposed into complementary temporal features such as dwell time and transition dynamics. Occurrences were computed by mapping the frames assigned to each μCAP back onto individual time series. They reflect the total number of time points during which a participant’s brain activity aligns with a given μCAP, while the corresponding subregion within the FFG seed is active. To test for the sex-by-diagnosis interaction in μCAP expression, we fit separate ordinary least-squares regression (OLS) models for each μCAP’s normalised occurrence count including diagnosis, sex, and their interaction as predictors, while controlling for age, meanFD, and site (i.e., covariates). P-values were corrected for multiple comparisons using FDR correction. Follow-up simple effects were evaluated using planned contrasts computed from the fitted regression models.

For μCAPs showing a significant sex-by-diagnosis interaction in normalised occurrence frequency, we further examined more fine-grained temporal properties, such as dwell time (i.e., average duration of uninterrupted expression) and transition probabilities to and from the significant μCAP. Differences in dwell time were analyzed using OLS regression models with the same predictors and covariates as above. To assess group differences in transition probabilities between μCAP states, we fit binomial generalized linear models (GLMs) for each transition, modeling the number of transitions from state *i* to state *j* as a function of diagnosis, sex, their interaction, and covariates, with the total number of transitions from state *i* as the binomial denominator. FDR correction was applied across all tested transitions involving the relevant μCAP.

Next, we examined the significant μCAP’s associations with core clinical measures related to social functioning and face processing within autistic individuals (for details, see Methods in Supplement). A linear regression model was fit with clinical scores as the dependent variable and the normalised occurrence as the main predictor, including an interaction term with sex and covariates.

Finally, we investigated whether the μCAP showing a significant group-by-sex interaction in occurrence also differed in its spatial expression, specifically in terms of its extent. To this end, we examined group-by-sex effects on its spatial extent (i.e., size). We first computed the mean group-level μCAP map across all individuals and 90th percentile thresholds were derived from the distribution of positive voxel values. The derived threshold was then applied to each individual μCAP map. Next, we computed the number of suprathreshold voxels in the individual μCAP maps and the individual voxel counts were normalised by total brain volume to account for interindividual differences in brain size. Subjects with zero suprathreshold voxels were excluded from that analysis, as these represent absence of μCAP activation at that threshold (N=2). An OLS model was fitted with normalised voxel count as the dependent variable and diagnosis, sex, and their interaction as fixed effects, while controlling for the same covariates as above. Analyses were repeated across multiple percentile thresholds (85–99%) to test for robustness of the result.

### Sensitivity Analyses

To ensure that results were not confounded by head motion, we removed participants with the highest motion values (N=17) until group differences in meanFD across diagnosis and sex were no longer significant (p>0.1). All analyses were repeated in the reduced, motion balanced sub-sample. Given differences in intellectual functioning across the groups, we also reran all analyses including full-scale IQ (FIQ) as additional covariate. For details, see the Supplement.

## Results

### Sample Characteristics

The final sample comprised 514 individuals (208 autistic males, 78 autistic females, 146 non-autistic males, 82 non-autistic females) between 6 and 30 years of age. All individuals were matched for age. Autistic individuals had lower measures of intellectual functioning than non-autistic individuals and autistic males had higher autistic symptom severity than autistic females. Demographic and clinical characteristics and group comparisons are listed in Table 1. For an overview of medication use, see Table S4.

**Table 1.**

| Variable | ASC M |  |  | ASC F |  |  | NAI M |  |  | NAI F |  |  | stats | posthoc |
| --- | --- | --- | --- | --- | --- | --- | --- | --- | --- | --- | --- | --- | --- | --- |
|  | Mean | SD | Range | Mean | SD | Range | Mean | SD | Range | Mean | SD | Range |  |  |
| Age | 17.04 | 5.27 | 7.48-30.31 | 17.73 | 5.91 | 7.88-30.26 | 17.24 | 5.48 | 7.57-30.95 | 17.24 | 5.46 | 6.89-28.5 | $F=0.29, p=0.83$ | n.s. |
| FIQ | 101 | 9.12 | 59-148 | 97 | 17.79 | 61-131 | 106.47 | 15.71 | 51-142 | 103 | 19.18 | 50-141 | $F=4.89, p=0.002$ | ASC M < NAI M; ASC F < NAI M & NAI F |
| VIQ | 100 | 19.19 | 52-160 | 98 | 17.47 | 51-136 | 104.78 | 16.48 | 51-142 | 104 | 19.91 | 51-147 | $F=3.66, p=0.01$ | ASC M < NAI M; ASC F < NAI M & NAI F |
| PIQ | 103 | 21.46 | 44-150 | 97 | 19.43 | 55-130 | 107.22 | 17.89 | 46-147 | 102 | 19.6 | 49-139 | $F=4.49, p=0.004$ | ASC M > ASD F; ASC M < NAI M; ASC F < NAI M |
| ADI Social | 16.78 | 6.32 | 1.0-28.0 | 15.58 | 7.17 | 1.0-29.0 | - | - | - | - | - | - | $p=0.17$ | n.s. |
| ADI Com | 13.5 | 5.52 | 0.0-26.0 | 12.32 | 5.24 | 0.0-24.0 | - | - | - | - | - | - | $p=0.1$ | n.s. |
| ADI RRB | 4.27 | 2.59 | 0.0-12.0 | 3.85 | 2.46 | 0.0-10.0 | - | - | - | - | - | - | $p=0.22$ | n.s. |
| ADOS CSS | 5.54 | 2.7 | 1.0-10.0 | 4.33 | 2.47 | 1.0-9.0 | - | - | - | - | - | - | $p=0.0007$ | ASC M > ASC F |
| ADOS SA CSS | 6.22 | 2.52 | 1.0-10.0 | 5.29 | 2.53 | 1.0-10.0 | - | - | - | - | - | - | $p=0.006$ | ASC M > ASC F |
| ADOS RRB CSS | 4.76 | 2.58 | 1.0-10.0 | 3.92 | 2.49 | 1.0-9.0 | - | - | - | - | - | - | $p=0.015$ | ASC M > ASC F |
|  | Median | SD | Range | Median | SD | Range | Median | SD | Range | Median | SD | Range |  |  |
| meanFD | 0.08 | 0.08 | 0.02-0.45 | 0.07 | 0.08 | 0.02-0.5 | 0.07 | 0.07 | 0.03-0.4 | 0.06 | 0.05 | 0.02-0.29 | $\chi^2=13.35, p=0.004$ | ASC M > NAI M & NAI F |
| RMET | 64.29 | 14.21 | 22.58-91.67 | 69.44 | 14.1 | 19.44-94.44 | - | - | - | - | - | - | $W=6877, p=0.05$ | ASC M < ASC F |

Abbreviations: ASC=autism spectrum condition; NAI=non-autistic individuals; FIQ=full-scale IQ; VIQ=verbal IQ; PIQ=performance IQ; ADI=Autism Diagnostic Interview; ADOS=Autism Diagnosis Observation Schedule; FD=framewise displacement; RMET=Reading the Mind in The Eyes Test (accuracy)

### μCAP Networks

To characterize the dynamic large-scale network engagement of the FFG, we identified recurring μCAPs. The optimal number of μCAPs was six based on a local drop in the maximum cluster assignment distance (Figure S2), indicating that the FFG dynamically engaged with six distinct large-scale networks (Figure 2A–F). Overlap between each μCAP and well-characterized resting-state systems (Figure 2H) is summarized in Table S5, reflecting partial engagement across multiple systems. The six μCAPs were functionally annotated according to their greatest spatial overlap with canonical Yeo networks (Figure 2G). Specifically, the μCAPs aligned as follows: the visual central network (μCAP1) involving occipital parts of the FFG, lingual gyrus, planum temporale, intra- and supracalcarine cortex and cuneal cortex; the ventral attention network (μCAP2) involving central opercular cortex, supplementary motor cortex, parietal operculum, insula and supramarginal gyrus; the default mode network - C (μCAP3) involving posterior cingulate gyrus, posterior parahippocampal gyrus, precuneus, and angular gyrus; the visual peripheral network (μCAP4) involving intracalcarine cortex, occipital parts of the FFG, supracalcarine cortex, lingual gyrus, cuneal cortex and occipital pole; the dorsal attention network (μCAP5) involving occipital parts of the FFG, inferior and superior lateral occipital cortex, inferior and middle temporal gyrus (temporo-occipital part) and superior parietal lobule; the default mode network - B (μCAP6) involving posterior temporal fusiform cortex, anterior and posterior parahippocampal gyrus, temporal occipital fusiform cortex, posterior inferior temporal gyrus (temporo-occipital part) and lingual gyrus. For further details, see Figure S3 and Table S5.

We next examined how these large-scale networks were spatially organised within the FFG. Results showed that each of the identified large-scale brain networks was associated with a specific data-driven subregion of FFG (Figure 3A, Figure S3). The visual peripheral network was linked to the most posterior portion of the FFG, while the DMN was linked to the most anterior portions of the FFG. This posterior-to-anterior functional gradient was further supported by the distinct overlap of the FFG subregions with the VIS atlas (Figure 3E). More posterior μCAPs (e.g., visual peripheral-related μCAP4 and dorsal attention-related μCAP5) were associated with early retinotopic and low-level visual areas such as V3v and character-related patches, while more anterior μCAPs (e.g., DMN-related μCAP3 and μCAP6) aligned with higher-order regions such as mFus and pFus (Figure 3B-D). As further confirmed by connectopic mapping analyses, functional connectivity gradients within the FFG revealed a consistent low-to-high organizational axis across hemispheres (Figure 4A). When μCAPs were projected onto this gradient, they exhibited distinct and ordered preferences along the axis, with some μCAPs preferentially occupying lower (e.g., visual peripheral-related μCAP4), others mid-range (e.g., visual central-related μCAP1), and others higher gradient bins (e.g., DMN-related μCAP3) (Figure 4B-C).

**Figure 4.**
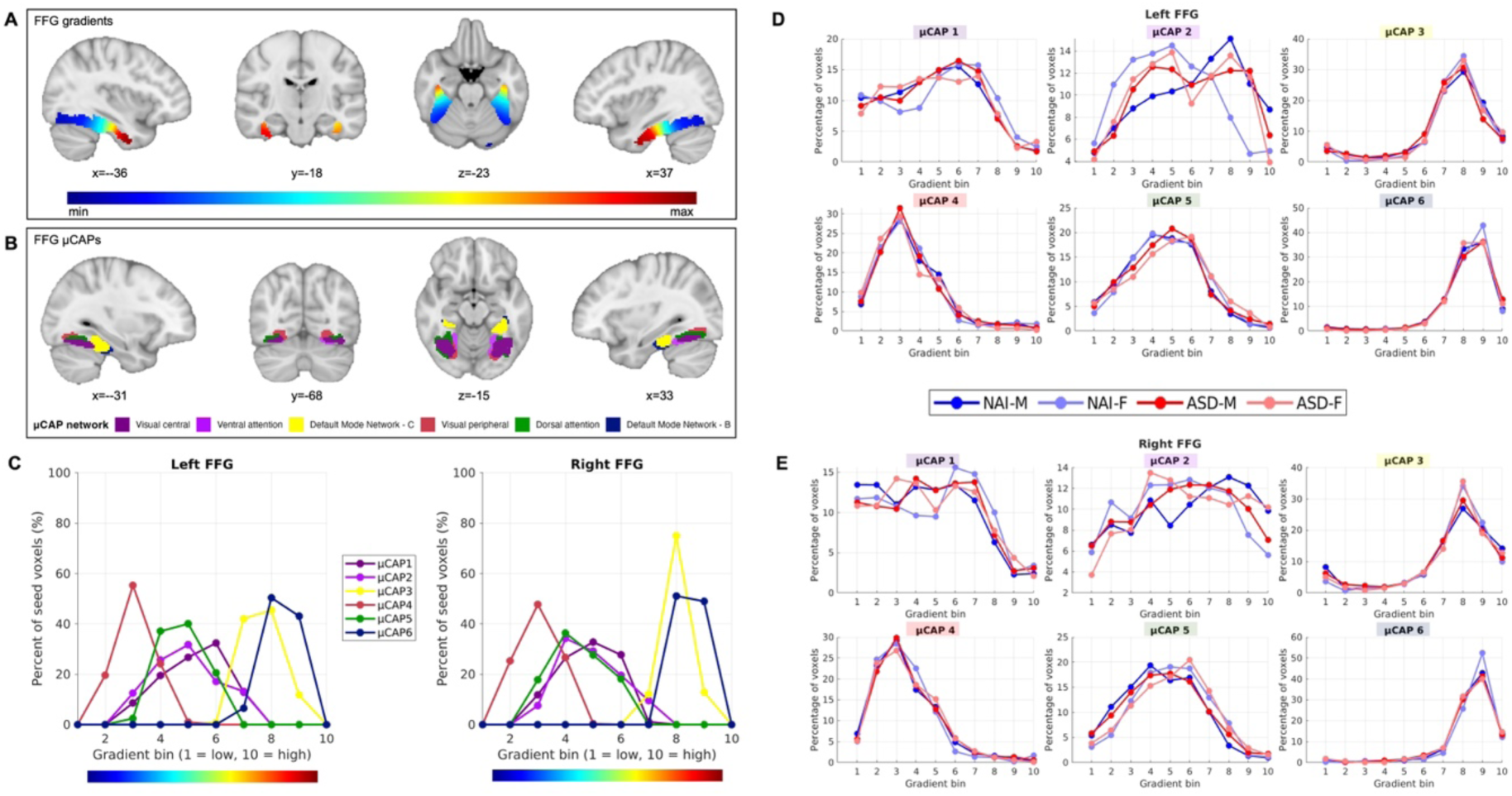
FFG Enrichment Profiles. (**A**) Group-average functional connectivity gradients of the left and right FFG, illustrating a principal axis of intrinsic organization from lower-level visual to higher-order associative regions. (**B**) Spatial maps of FFG μCAP seed parcels overlaid on anatomical templates, color-coded by large-scale functional network affiliation with Yeo atlas. (**C**) Distribution of μCAP voxels across gradient bins in the left and right FFG, expressed as the percentage of voxels within each μCAP falling into successive gradient bins (1=low, 10=high). μCAPs exhibit distinct and ordered enrichment profiles along the gradient, with consistent patterns observed across hemispheres. (**D-E**) Subject-level enrichment profiles for all six μCAPs for both the left (**D**) and right (**E**) hemispheres.

We next examined whether μCAP enrichment along the FFG functional gradient differed as a function of diagnosis and sex to test whether group differences reflected systematic shifts or dispersion along the FFG’s functional hierarchy. For the ventral attention-related μCAP2, there was a significant interaction between gradient bin, diagnosis, and sex (right FFG: F(1, 5096)=10.07, *p*=0.002, *pFDR*=0.009; left FFG: F(1, 5079)=11.88, *p*=0.0006, *pFDR*=0.003), indicating that sex-dependent differences in μCAP2 enrichment along the FFG functional gradient were differentially expressed in autistic and non-autistic individuals. Follow-up bin-specific contrasts revealed that sex differences were present in non-autistic individuals at lower and higher gradient bins, whereas no reliable sex or diagnosis differences were observed within autistic individuals (see Table S6). More specifically, results indicated that non-autistic females showed a more early-gradient enrichment, whereas non-autistic males exhibited relatively greater enrichment in higher gradient bins. In contrast, both autistic males and females displayed more diffuse distributions across gradient bins, indicating less distinct localization of μCAP expression along the functional hierarchy (Figure 4D-E).

### Temporal, Clinical and Spatial Characterisation of μCAPs

We next characterised the temporal features of μCAP expression, focusing on potential sex-and diagnosis-related differences. Among all six μCAPs, there were no significant main effects of diagnosis and sex, however, there was a significant sex-by-diagnosis interaction for the occurrences of the DMN-related μCAP 3 (β=0.003, Standard Error [SE]=0.001, 95% CI [0.001, 0.005], *t*=2.88, *p*=0.004, *pFDR*=0.025) (Figure 5A and Table S7). Post-hoc tests indicated that non-autistic females showed a higher occurrence of this μCAP than non-autistic males (β=-0.006, SE=0.002, 95% CI [-0.009,0.002], *t*=-3.7, *p*<0.001, *pFDR*<0.001), while no sex difference was observed among autistic individuals (β<0.001, SE=0.002, 95 % CI [-0.003, 0.004], *t*=0.3, *p*=0.77, *pFDR*=0.77; see Figure 5A).

**Figure 5.**
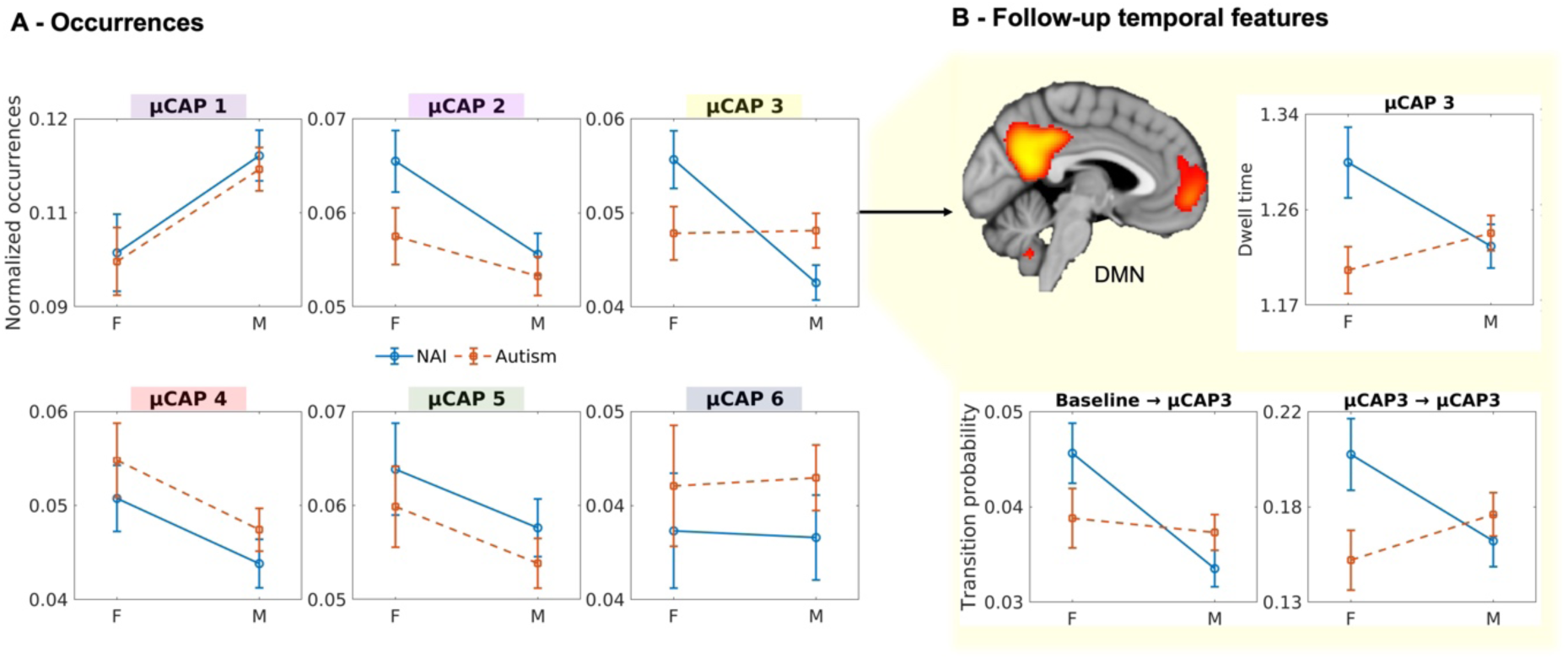
Temporal Features. (**A**) Sex-by-diagnosis interactions for the normalised occurrences across all six μCAPs. There was a significant interaction for the default mode network-related μCAP 3 with non-autistic females showing higher frequency compared to non-autistic males, while this sex difference was absent in autistic individuals. (**B**) A similar sex-by-diagnosis interaction pattern emerged for the follow-up temporal features for the significant μCAP 3 including dwell time and transition probabilities.

We next further characterized this μCAP 3 in terms of finer-grained temporal dynamics, including dwell time and transition probabilities. Results revealed a significant sex-by-diagnosis interaction for dwell time (β=0.027, 95% CI [0.005, 0.048], SE=0.011, *t*=2.43, *p*=0.015) with non-autistic females showing longer average time spent in μCAP 3 than non-autistic males (β=-0.04, 95% CI [-0.06, -0.005], SE=0.015, *t*=-2.29, *p*=0.02), and autistic males and females showing no sex difference (β=0.018, 95% CI [-0.013, 0.047], SE=0.015, *t*=1.13, *p*=0.26) (Figure 5B). Furthermore, we observed significant sex-by-diagnosis interactions in the transition probabilities both from the baseline state to μCAP 3 (Odds ratio [OR]=1.27, 95 % CI [1.08, 1.49], *Z*=2.87, *p*=0.0041, *p*FDR=0.029) and in the probability of remaining in μCAP 3 across consecutive time points (OR=1.51, 95% CI [1.14, 2.01], *Z*=2.84, *p*=0.0045, *p*FDR=0.029) (Figure 5B). Post hoc comparisons revealed that non-autistic females were more likely than non-autistic males to transition into (OR=0.77, 95% CI [0.68, 0.87], *Z*=-4.35, *p*<0.001, *p*FDR<0.001) and persist in μCAP 3 (OR=0.8, 95% CI [0.66, 0.98], *Z*=-2.16, *p* = 0.03, *p*FDR=0.22), whereas no significant sex differences were observed among autistic individuals (OR=0.98, 95% CI [0.87, 1.1], *Z*=-0.35, *p*=0.73, *pFDR*=0.94 and OR=1.22, 95% CI [0.99, 1.49], *Z*=1.86, *p*=0.06, *pFDR*=0.52, respectively). All results remained significant in a motion-balanced sample and when additionally controlling for FIQ (see Results in Supplement).

To assess clinical relevance, we examined associations between μCAP 3 occurrence and measures of social functioning. There was a nominally significant interaction between sex and normalised occurrence counts of μCAP 3 on the ADI-social (β=1.85, SE=0.87, 95% CI [0.131, 3.568], *t*=2.12, *p*=0.03), suggesting a potential sex-dependent association between μCAP 3 occurrences and social features in autistic individuals, however, this effect did not survive FDR correction (*pFDR*=0.1) (Figure S4). This interaction was not significant for ADOS social (β=0.5, SE=0.35, 95% CI [-0.19, 1.18], *t*=1.44, *p*=0.15, *pFDR*=0.23) or performance on the Reading the Mind in the Eyes Test (β=-0.03, SE=0.15, 95% CI [-0.33, 0.27], *t*=-0.18, *p*=0.86, *pFDR*=0.86).

When examining spatial extent of μCAP 3, we observed a significant sex-by-diagnosis interaction (β=−0.001, SE<0.001, 95% CI [-0.003, -0.0005], *t*=−2.03, *p*=0.043) using a 90^th^percentile threshold to define suprathreshold voxels. The result indicated that non-autistic males showed a larger μCAP 3 extent than non-autistic females (β=−0.003, SE=0.001, 95% CI [-0.005, -0.0008], *t*=−2.7, *p*=0.007), whereas the opposite pattern was observed among autistic individuals (β=-0.006, SE=0.001, 95% CI [-0.008, -0.004], *t*=-5.7, *p*<0.001) (Figure S5B). When repeating analysis across a range of percentile thresholds (85–95%), the sex-by-diagnosis interaction effect remained comparable across thresholds (Figure S5A).

## Discussion

In this study, we investigated the neural organization of face-processing systems, a domain in which autistic individuals commonly experience difficulties. We focused on the FFG, a central hub of the face-processing network, and examined how its intrinsic functional coupling with large-scale brain networks varies across time. Using the micro co-activation patterns (μCAPs) approach, we characterized the dynamic configurations through which the FFG engages with large-scale brain networks, specifically testing how these patterns differ between autistic and non-autistic males and females. Whereas prior work has primarily characterized autism- and sex-related differences in large networks in terms of static functional connectivity, the present results demonstrate that such differences are also reflected in the temporal dynamics with which DMN-related states are entered and sustained. These results highlight that sex- and diagnosis-related variation extends beyond average connectivity strength to the dynamic organization of transient network configurations.

We identified six μCAPs mapping onto large-scale canonical brain networks, including peripheral and central visual networks, dorsal and ventral attention networks, and different portions of the DMN. These networks were each associated with a discrete subregion within the FFG which exhibited a posterior-to-anterior functional organization, whereby lower-level visual subareas mapped to posterior regions and higher-order subareas, including two subdivisions of the DMN, mapped to anterior regions. This organization of μCAPs within the FFG aligns with its known topographical functional architecture [44–46], recapitulating the expected posterior-to-anterior transition from lower-level visual processing to higher-level representations, and providing convergent support for the biological validity of the seed definition. Our findings further extend prior work by showing that the FFG contains multiple subdivisions linked to both perceptual and socio-cognitive processes that are not static, but dynamically functionally coupled with distinct large-scale systems.

Notably, this functional hierarchical alignment was largely consistent across diagnostic and sex groups. Only for μCAP 2 related to ventral attention non-autistic males and females showed different positions along the FFG gradient. Prior research has shown that both interindividual variability [78] and sex differences [79–81] in the spatial organization of functional networks are most pronounced in association networks, including the ventral attention network and DMN. Consistent with this, we observed sex-dependent differences in the spatial topography of ventral attention-related μCAPs within the FFG in non-autistic individuals, suggesting differences in functional specialization [82] and organizational strategies between the sexes [83]. In contrast, this topographical segregation appeared attenuated in autistic individuals, who engaged a broader, more diffuse distribution of ventral attention–related μCAPs across the FFG. As the ventral attention network supports bottom-up orienting toward salient visual information [84], different gradient embedding may reflect differences in how salient facial features are processed. This aligns with prior reports that autistic individuals allocate visual attention differently during face perception, for example showing reduced attention to the eye region and greater focus on other facial features [85–87].

When examining the temporal dynamics of identified μCAPs, only the DMN-C μCAP 3 linked to higher-order anterior FFG regions, showed sex-by-diagnosis differences across multiple temporal measures. Non-autistic females exhibited more frequent engagement of this μCAP than non-autistic males, accompanied by longer dwell times and a higher likelihood of transitioning into and remaining in this state, consistent with more sustained coupling between anterior FFG regions and the DMN. This pattern aligns with well-established sex differences in DMN functional connectivity, with non-autistic females typically showing stronger DMN connectivity than non-autistic males [23, 88–90]. In contrast, these sex differences were markedly attenuated in autism, with autistic males and females engaging this network equally often, maintaining it for similar durations, and transitioning into and persisting in this state to a similar extent. The DMN has been repeatedly implicated in autism [91], and pronounced sex differences have been reported, with both autistic males and females and non-autistic males exhibiting hypo-connectivity compared to non-autistic females [22, 23, 92]. Notably, this established pattern closely parallels the present temporal findings, suggesting that autism is associated with a reduction of typical sex differentiation in the engagement of anterior FFG–DMN circuitry. Given the central role of the DMN in mentalizing and social-cognitive processes [93], these results point to differences in the recruitment of higher-order socio-cognitive networks during face processing rather than alterations in early perceptual mechanisms. This pattern is consistent with both neuroimaging and behavioural evidence of reduced sex differences in social-emotional domains [22, 23, 94, 95]. Together, these findings thus suggest that dynamic interactions between higher-order FFG subdivisions and the DMN are important for understanding sex-related variability in autism.

With regards to network size, non-autistic males showed a larger spatial extent of μCAP 3 than non-autistic females, suggesting that when the anterior FFG–DMN configuration is expressed, non-autistic males engage a more spatially diffuse co-activation pattern. This is consistent with prior reports of greater within-network dispersion of DMN regions in non-autistic males compared to non-autistic females [81]. In autistic individuals, the opposite pattern was observed, with autistic females having larger spatial extent compared to autistic males. This reversal of sex-typical patterns indicates that autism is associated with a reorganization of the spatial architecture through which anterior FFG–DMN coupling is achieved in non-autistic individuals. Autistic females may recruit a broader, more diffuse network to support functions typically mediated by this circuit. A less spatially constrained integration between FFG and higher-order association cortex may potentially reflect reduced functional specialization [96, 97] or compensatory redistribution of social-perceptual processing demands. These findings indicate that sex differences in functional DMN coupling are not merely attenuated in autism, but also spatially reorganized.

Prior work has linked differences in temporal network dynamics to autistic traits and symptom severity. In the present study, we observed a trend-level association between μCAP3 occurrence and social difficulties that differed across autistic males and females, suggesting that dynamic features of network organization may relate to variability in social functioning. More broadly, recent work indicates that temporal properties of brain network organization may serve as clinically informative markers, with distinct alterations in co-activation pattern dynamics observed across different psychiatric conditions [98]. Our findings further suggest that sex-related variability in autism may partly be related to differences in the dynamic recruitment of higher-order socio-cognitive networks, rather than uniform alterations in perceptual regions. In particular, the attenuation and reorganization of typical sex differences in anterior FFG–DMN dynamics highlight the importance of considering sex as a stratification factor in both research and intervention design. Dynamic network features may help identify subgroups characterized by distinct patterns of socio-cognitive network engagement, opening avenues for more personalized approaches. Although preliminary, these findings raise the possibility that interventions targeting socio-cognitive networks could be refined by accounting for individual differences in dynamic network organization. For example, approaches such as neuromodulation or social-cognitive training may benefit from tailoring strategies depending on whether their baseline dynamic coupling profiles differ or alterations reflect reduced sustained engagement of higher-order networks or differences in perceptual–attentional integration within the FFG. Longitudinal studies will be essential to determine whether these dynamic signatures predict treatment response, track developmental trajectories, or serve as stratification markers for sex-informed intervention strategies.

It remains to be established whether genetic and hormonal mechanisms that typically contribute to sexual differentiation in non-autistic individuals may influence these functional patterns and how these mechanisms may be different or reorganized in autism. Previous studies have shown that brain structure and function in autism can show a greater resemblance with the non-autistic male pattern [10, 20, 23, 99] which was also the case across examined temporal features. Interestingly, our prior work shows that greater resemblance to the non-autistic male pattern in autistic females was associated with poorer mentalizing and face processing performance [10]. Furthermore, variation in prenatal androgens during mid-gestation exerts sex-differential effects on network organization relevant to social cognition with elevated levels predicting reduced functional connectivity within the DMN in non-autistic males but not in non-autistic females [100]. These findings suggest that sex-differential neurodevelopmental mechanisms may shape functional brain organization in autism, particularly within networks supporting social cognition. Whether the same mechanisms extend to dynamic network states and functionally distinct subregions of social processing areas remains to be determined.

Finally, we focused on resting-state dynamics, as μCAP analyses are specifically designed to capture intrinsically emerging network configurations rather than stimulus-locked responses. Resting-state data allow characterization of intrinsic, task-independent network dynamics, which may reflect more stable, trait-like aspects of brain organization relevant to autism. In addition, task paradigms introduce structured temporal constraints and performance-related variability (e.g., differences in engagement or strategy use) that may limit the ability to isolate spontaneous co-activation patterns and complicate the interpretation of group- and sex-related differences in large and heterogeneous samples. Nevertheless, whether similar dynamic signatures are expressed during face processing tasks remains an important question for future work. Sex was modeled as a binary variable, and we lacked measures of pubertal development, endocrine status, gender-related experiences and gender identity. Therefore, we cannot disentangle biological from gendered developmental contributions to the observed sex-differential patterns. Finally, the cross-sectional design limits inferences regarding developmental trajectories of sex-differential dynamic organization. Given the wide age range of the sample, future longitudinal work will be essential to capture how sex-differential FFG–DMN dynamics emerge and evolve across development.

## Conclusion

In summary, we report both attenuation and reversal of typically occurring sex differences in the functional configuration and spatial extent of anterior FFG–DMN circuitry. Extending beyond static functional connectivity measures, we demonstrate that these differences manifest not only in spatial network organization but also in the temporal dynamics with which specific FFG–DMN configurations are entered and maintained. These findings align with the notion that autism affects the processes that typically contribute to sexual differentiation of social-cognitive functions and highlight the importance of considering sex as a biological variable when examining neural heterogeneity in autism.

## Data Availability

All data produced in the present study are available upon reasonable request to the authors

## Acknowledgements

We thank all participants and their families for participating in the studies that contribute to the datasets used in this research. We also gratefully acknowledge the contributions of all members of the EU-AIMS/AIMS-2-TRIALS LEAP group: Jumana Ahmad, Sara Ambrosino, Bonnie Auyeung, Sarah Baumeister, Sven Bölte, Carsten Bours, Michael Brammer, Daniel Brandeis, Claudia Brogna, Yvette de Bruijn, Bhismadev Chakrabarti, Ineke Cornelissen, Daisy Crawley, Guillaume Dumas, Jessica Faulkner, Vincent Frouin, Pilar Garcés, David Goyard, Lindsay Ham, Hannah Hayward, Joerg Hipp, Mark H. Johnson, Emily J.H. Jones, Xavier Liogier D’ardhuy, David J. Lythgoe, René Mandl, Luke Mason, Andreas Meyer-Lindenberg, Nico Mueller, Bethany Oakley, Laurence O’Dwyer, Bob Oranje, Gahan Pandina, Antonio M. Persico, Barbara Ruggeri, Amber Ruigrok, Jessica Sabet, Roberto Sacco, Antonia San José Cáceres, Emily Simonoff, Will Spooren, Roberto Toro, Heike Tost, Jack Waldman, Steve C.R. Williams, Caroline Wooldridge, and Marcel P. Zwiers. This project has received funding from the Innovative Medicines Initiative 2 Joint Undertaking under grant agreement No 115300 (for EU-AIMS) and No 777394 (for AIMS-2-TRIALS). This Joint Undertaking receives support from the European Union’s Horizon 2020 research and innovation programme and EFPIA and AUTISM SPEAKS, Autistica, SFARI. Any views expressed are those of the author(s) and not necessarily those of the funders (IHI-JU2). We also gratefully acknowledge the contributions of all members of the APEX (Autism and Prenatal Sex Differences) consortium: Deep Adhya, Carrie Allison, Bonnie Ayeung, Rosie Bamford, Simon Baron-Cohen, Richard Bethlehem, Tal Biron-Shental, Graham Burton, Wendy Cowell, Jonathan Davies, Joanna Davis, Dorothea Floris, Alice Franklin, Lidia Gabis, Daniel Geschwind, David M. Greenberg, Yuanjun Gu, Alexandra Havdahl, Alexander Heazell, Rosemary Holt, Matthew Hurles, Yumnah Khan, Meng-Chuan Lai, Madeline Lancaster, Michael Lombardo, Hilary Martin, Jose Gonzalez Martinez, Jonathan Mill, Mahmoud Musa, Kathy Niakan, Adam Pavlinek, Lucia Dutan Polit, Marcin Radecki, David Rowitch, Jenifer Sakai, Laura Sichlinger, Deepak Srivastava, Alexandros Tsompanidis, Florina Uzefovsky, Varun Warrier, Elizabeth Weir, Xinhe Zhang. This work was also supported by the Netherlands Organization for Scientific Research through Vidi grants (Grant No. 864.12.003 [to CFB]; from the FP7 (Grant No 278948 (TACTICS); and from the European Community’s Horizon 2020 Programme (H2020/2014-2020) (Grant Nos. 643051 [MiND], 642996 (BRAINVIEW) and 847818 (CANDY). This work received funding from the Wellcome Trust UK Strategic Award (Award No. 098369/Z/12/Z) and from the National Institute for Health Research Maudsley Biomedical Research Centre (to DM). DLF is supported by the UZH Postdoc Grant, grant no. [FK-23-085]. RH received funding from SFARI GAIINS (grant number 10039678). SBC received funding from the Wellcome Trust 214322\Z\18\Z. SBC also received funding from the Innovative Medicines Initiative 2 Joint Undertaking under grant agreement No 777394 for the project AIMS-2-TRIALS. The University of Cambridge Autism Research Centre received funding for this work from the Autism Research Trust, whose legacy work is now managed by Autism Action. SBC also received funding from SFARI, the Templeton World Charitable Fund and the MRC. He is grateful to Cambridge University Development and Alumni Relations (CUDAR) for anonymous donations. All research at the Department of Psychiatry in the University of Cambridge is supported by the NIHR Cambridge Biomedical Research Centre (NIHR203312) and the NIHR Applied Research Collaboration East of England. The views expressed are those of the author(s) and not necessarily those of the NIHR or the Department of Health and Social Care. BHV is supported by funding from the UZH Postdoc Grant, grant no. [FK-23-086].

