## Supplementary material for "Attenuation of typical sex differences in the time-resolved functional connectivity of the fusiform gyrus in autism": Supplment

### Participants, study design and exclusion criteria

We used participants from the EU-AIMS Longitudinal European Autism Project (LEAP). Full details of the LEAP study design can be found in ^1,2^. The study was approved by local research ethics committees at each site (via the Integrated Research Application System [IRAS], UK; see Table S1), and written informed consent was obtained from all participants or their legal guardians (for participants <18 years). All autistic participants had an existing clinical diagnosis of autism according to DSM-IV^3^, DSM-IV-TR^4^, DSM-5^5^ or ICD-10^6^ criteria. In the autism group, diagnosis was confirmed using the combined information from gold-standard diagnostic instruments, the Autism Diagnostic Interview-Revised^7^ (ADI-R) and the Autism Diagnostic Observation Schedule (ADOS)^8^. Appropriate to a multi-centre study, quality control procedures were in place around training and data collection/entry. Cross-site training sessions for collecting clinical data were put in place, the ADOS and ADI-R were administered and scored by qualified/certified personnel and the study was regularly monitored according to Good Clinical Practice (GCP) standards. Participants underwent comprehensive clinical, cognitive, and MRI assessment at one of six collaborating sites: the Institute of Psychiatry, Psychology and Neurosciences, King’s College London (KCL), London, United Kingdom; Autism Research Centre at the University of Cambridge, Cambridge, United Kingdom; Radboud University Nijmegen Medical Centre, Nijmegen, the Netherlands; University Medical Centre Utrecht, Utrecht, the Netherlands; Central Institute of Mental Health, Mannheim, Germany; and University Campus Bio-Medico, Rome, Italy. Exclusion criteria included the presence of any MRI contraindications (e.g., metal implants, braces, claustrophobia) or failure to give informed written consent to MRI scanning, as well as significant hearing or visual impairments not corrected by glasses or hearing aids. In addition to general exclusion criteria, we also excluded individuals based on the imaging data. Resting-state fMRI data (rs-fMRI) was available for 615 subjects in five out of the six acquisition sites (not collected for the Rome site). One person was excluded due to consent withdrawal. Images with clinically (mostly) non-significant atypicalities (N=18) and excessive head motion (N=21) were excluded. Furthermore, scans with extreme values (N=19; values >4SD), with less than 180 volumes (i.e., less than 90% of rs-fMRI scan completed; N=5), low full-brain coverage (N=19), a TR different than the standard TR =2.3 s (N=1), excessive head motion during the rs-fMRI scan (N=12 with mean root-mean-square of the framewise displacement [meanFD] >0.7 and N= 8 with maxFD >7.6 [i.e., motion of more than 2 voxels]), with more than 25% scrubbed volumes (N=6) and low fusiform gyrus (FFG) coverage (N=1) were excluded. For a detailed flowchart on exclusions, see Figure S1. Applying all exclusion criteria resulted in a sample of 514 individuals (208 autistic males, 78 autistic females, 146 non-autistic males, 82 non-autistic females) between 6 and 30 years of age. For further details, see Table 1.

### Demographic, clinical, and cognitive measures

#### Intellectual functioning

General intellectual abilities were assessed using the Wechsler Abbreviated Scales of Intelligence-Second Edition^9^ (WASI-II), or if unavailable the Wechsler Intelligence Scale for Children-III/IV^10,11^ (WISC-III/IV) for children or Wechsler Adult Intelligence Scale-III/IV^12,13^ (WAIS-III/IV) for adults. Standardized estimates of verbal IQ (VIQ), performance IQ (PIQ), and full-scale IQ (FIQ) were derived using IQ norms with mean=100 and SD=±15.

#### ADOS

The Autism Diagnostic Observation Schedule^8^ (ADOS-G) was used to measure the impact of current, clinically observed core symptoms of autism. Based on ADOS-2 algorithm totals^14,15^, we report ADOS-2 Calibrated Severity Score (CSS) for ‘Social Affect’ indexing social-communication difficulties and ‘RRBs’ indexing restricted and repetitive behaviours. The CSS scores range from 1 to 10, with higher scores indicating more severe autism symptom severity.

#### ADI-R

The Autism Diagnostic Interview-Revised^7^ (ADI-R) is a structured parent interview completed by parents or caregivers of participants with autism. Algorithm scores were derived from current and historical symptom information for the domains of Reciprocal Social Interaction, Communication, and Restricted, Repetitive and Stereotyped Behaviours and Interests.

#### The Reading the Mind in the Eyes Test

The Reading the Mind in the Eyes test^16^ (RMET) asks participants to identify complex emotions and mental states based only on the eye region of a face. Depending on their age (adults: 18-30, adolescents: 12-17, children: 6-11) and ability level, participants received either an adult (36 items), adolescent (31 items) or child (28 items) version of the test. Percentage of correct answers was used as the outcome variable. Before merging the three different versions across all subjects, each age-related version was z-standardized.

#### Missing clinical data

To address missing clinical data and avoid further reducing sample size, we used imputed clinical data^17^, as in previous work with this dataset^18,19^. The imputation procedure considered the potential non-randomness of missing data, and therefore developed quantitative measures to assess the quality of the imputations, and finally imputed data adopting a nonparametric tree regression model embedded in an iterative round-robin schedule. The exact procedure within this dataset has been published^17^.

### MRI data acquisition

MRI data were acquired on 3T scanners: General Electric MR750 (GE Medical Systems, Milwaukee, WI, USA) at Institute of Psychiatry, Psychology and Neuroscience, King’s College London, United Kingdom (KCL); Siemens Magnetom Skyra (Siemens, Erlangen, Germany) at Radboud University Nijmegen Medical Centre, the Netherlands (RUNMC); Siemens Magnetom Verio (Siemens, Erlangen, Germany) at Autism Research Centre at the University of Cambridge, United Kingdom (UCAM); Philips 3T Achieva (Philips Healthcare Systems, Best, The Netherlands) at University Medical Centre Utrecht, the Netherlands (UMCU); GE Medical Systems Signa HDxTt at the Rome University; and Siemens Magnetom Trio (Siemens, Erlangen, Germany) at Central Institute of Mental Health, Mannheim, Germany (CIMH). Procedures were undertaken to optimize the MRI sequences for the best scanner-specific options, and phantoms and travelling heads were employed to ensure standardization and quality assurance of the multi-site image-acquisition.

**T1-weighted images**: images were obtained using a 5.5 minute MPRAGE sequence (Nijmegen site example: TR=2300ms, TE=2.93ms, T1=900ms, voxel size=1.1x1.1x1.2mm, flip angle=9°, matrix size=256x256, FOV=270mm, 176 slices). Slight variations are present across centres, for details see Table S2.

**Resting-state fMRI**: An eight-to-ten minute rs-fMRI scan was acquired using a multi-echo planar imaging (ME-EPI) sequence developed by Kundu et al.^20^; TR=2300ms, TE~12ms, 31ms, and 48ms (slight variations are present across centres), flip angle=80°, matrix size=64x64, in-plane resolution=3.8mm, FOV=240mm, 33 axial slices, slice thickness/gap=3.8mm/0.4mm, volumes=200 (UMCU), 215 (KCL, CIMH), or 266 (RUNMC, UCAM). Participants were instructed to relax and fixate on a cross presented on the screen for the duration of the rs-fMRI scan. For further details, see Table S3.

### Resting-state fMRI data preprocessing

After combining the three rs-fMRI scan echoes using echo-time weighted averaging, the rs-fMRI data were preprocessed using a standard preprocessing pipeline that included tools from the FMRIB Software Library (FSL version 5.0.6; http://www.fmrib.ox.ac.uk/fsl). Preprocessing included removal of the first five volumes to allow for signal equilibration, primary head motion correction via realignment to the middle volume (MCFLIRT), grand mean scaling and spatial smoothing with a 6mm FWHM Gaussian kernel. Next, we thoroughly corrected for secondary head-motion related artifacts, by applying ICA-AROMA, an ICA-based method, which automatically detects and removes motion-related components from the data^21^. ICA-AROMA has been demonstrated to remove head motion-related artifacts with high accuracy while preserving signal of interest^21,22^. Finally, we applied nuisance regression to remove signal from white matter and cerebrospinal fluid, and a high-pass filter (0.01 Hz). The rs-fMRI images of each participant were coregistered to the participants' anatomical images via boundary-based registration implemented in FSL FLIRT^23^. The T1 images of each participant were registered to MNI152 standard space using 12-parameter affine transformation and refined using non-linear registration with FSL FNIRT (10mm warp, 2mm resampling resolution). Finally, we brought all participant-level rs-fMRI images to 2mm MNI152 standard space by applying the rs-fMRI to T1 and T1 to MNI152 transformations. All further analyses were conducted in MNI152 standard space.

### Region of interest – fusiform gyrus

The fusiform gyrus (FFG) served as the seed region in the subsequent co-activation pattern analysis. The FFG ROI was created by combining four regions of the Harvard-Oxford atlas (HOA) (fMRIB, Oxford, UK) (i.e., anterior and posterior divisions of the temporal fusiform cortex, temporal occipital fusiform cortex and occipital fusiform gyrus) for both the right and left hemisphere. Participants who had insufficient FFG coverage (i.e., less than 90%, N=1) were excluded. Based on the remaining participants, we reduced the FFG ROI to have 100% coverage across all individuals’ rs-fMRI scans. This made up 87.7% of the original left FFG ROI and 87.6% of the original right FFG ROI from the HOA.

### Co-activation pattern analysis

#### Overview

Co-activation pattern (CAP) analysis characterizes dynamic functional interactions between a seed region and distributed brain networks by decomposing fMRI data at the single-frame level. This approach aligns with models proposing discrete transitions between large-scale brain states and has been supported by multiple physiological validations^24,25^. In contrast to conventional static connectivity methods, which average across time, CAP analysis retains temporal resolution and allows overlapping spatial patterns of co-activation to emerge, rather than treating them as mutually exclusive. Moreover, unlike other dynamic functional connectivity approaches that operate on continuous time courses, CAPs use individual fMRI frames as the unit of analysis, making the method particularly sensitive to brief, recurring episodes of co-activation and their temporal variability^24,26^. CAPs analysis has been described in detail elsewhere^26,27^. Methodologically, CAPs are identified by first extracting and Z-scoring the mean BOLD time series from the seed ROI and selecting volumes exceeding a predefined threshold. These selected whole-brain frames across subjects are then submitted to k-means clustering, which partitions them into distinct co-activation patterns such that within-cluster similarity is maximized relative to between-cluster similarity. Each cluster centroid is converted into a spatial Z-map, reflecting the statistical deviation of voxelwise activation from zero. Further details on the clustering procedure are provided elsewhere^28^.

#### Steps

The μCAP algorithm was implemented as previously described in ^29^. In brief, the approach identifies a number (‘k’) of transient co-activation patterns of fMRI volumes based on activity within a seed region of interest (ROI), here the fusiform gyrus. At the first iteration, the mean BOLD signal of the seed voxels was used as the reference time course for frame selection. In subsequent iterations, seed time courses were recalculated by averaging fMRI volumes weighted by the current seed weight maps, allowing smaller subregions within the seed to contribute equally to subsequent frame selection. Frames were tagged as ‘selected’ when the Z-scored seed time course exceeded a threshold and only positive deviations (i.e., activation events) were retained, while deactivations were not considered here. The first iteration used a liberal threshold (Z > 0.5), followed by the conventional threshold (Z > 1.0) commonly applied in CAP studies^26,30^. Frames with excessive motion (FD > 0.5 mm) and their immediate neighbours (t−1, t+1) were excluded from selection (i.e., scrubbing). The retained frames were **projected to a low-dimensional principal-component (PC) space.** This step reduces voxel-level dimensionality and noise while preserving the dominant covariance structure among frames, which improves cluster stability and guards against overfitting. K-means clustering was then performed in this PC space with a correlation distance metric. The resulting cluster centroids were back-projected to voxel space to obtain μCAP maps, each representing a transient co-activation pattern. The centroid of each k-cluster represents a micro co-activation pattern (i.e., µCAP). Candidate solutions ranging from **k = 2 to 10** clusters were tested. The optimal number of clusters was chosen using a train–test stability procedure: clusters were defined on the training set, test frames were projected into the same PC space and assigned to training centroids to which they had the lowest distance. This distance was calculated as 1 – correlation to match the distance used within the k-means algorithm. The cluster with the largest assignment error (‘worst-fit’ cluster) was monitored across k. The optimal k was defined as the point where the worst-fit cluster showed a local drop in summed assignment distances across candidate solutions. Following clustering, each μCAP was restricted to the seed ROI, and its values were assigned to update the seed weights. A winner-takes-all strategy was applied within each voxel, assigning the highest μCAP value and setting others to zero. This updated seed weight map was then used for the next round of frame selection. Iterations continued until convergence, defined as the average cosine distance between matched μCAPs of successive iterations falling below 0.005. Matching was assessed using the Hungarian algorithm^31^. The initialization step with a uniform seed was not included in the convergence check. For further details, see ^29^ and Figure 1.

#### Occurrences

To quantify the frequency of μCAP expression over time, we computed subject-specific occurrence counts and fractions. For each participant and for each μCAP state, we counted the number of frames it was active, excluding all scrubbed frames. This resulted in a subject × state matrix of raw occurrence counts. To account for potential differences in the total number of non-scrubbed frames across participants, we also computed normalised occurrence fractions by dividing each state’s count by the total number of valid frames for that subject. This served as the primary feature of interest in analyses.

#### Follow-up temporal features

For μCAPs showing a significant sex-by-diagnosis interaction in normalised occurrence frequency, we further examined more fine-grained temporal properties, such as dwell time (i.e., average duration of uninterrupted expression) and transition probabilities to and from the significant μCAP.

##### Dwell Time

For each participant, we computed the mean dwell time for the previously significant μCAP. This was defined as the average number of consecutive time frames spent in the state before transitioning to another state. The state sequence was derived from the temporal clustering output, excluding scrubbed frames. The resulting dwell time reflects temporal stability or persistence of that μCAP.

##### Transition Probabilities

To quantify dynamic switching of the previously significant μCAP, we computed subject-specific transition probability matrices. Each participant’s framewise μCAP sequence was extracted from the time series. Transitions between consecutive valid frames (excluding any transitions involving scrubbed frames) were counted to form subject-level transition count matrices. These matrices were then row-normalised to yield transition probabilities, where each element represents the probability of transitioning from one state to another in the next time frame, conditional on the current state. Self-transitions were retained.

### Head motion artifact control

We applied a comprehensive procedure to minimize motion-related artifacts. At the preprocessing stage, primary motion correction was performed via realignment to the middle volume (MCFLIRT), followed by ICA-AROMA for data-driven removal of motion-related independent components. Participants with excessive head motion (> 3mm translation/rotation), as well as with high mean FD (> 0.7) and extreme maximum FD (> 7.6) were excluded. Frames with FD>0.5 mm and their immediate neighbors (t−1, t+1) were scrubbed prior to μCAP analysis. To further control for residual effects, all analyses included meanFD as a covariate. Furthermore, we conducted sensitivity analyses as shown below.

### Connectivity Gradients

Using the FFG ROI we estimated connection topographies^32^ separately for each individual and hemisphere using the *congrads* tool publicly available at <https://github.com/koenhaak/congrads>. This method computes for every voxel within the ROI the correlation between its voxel-wise time series and those of the rest of the cortex (based on a lossless singular value decomposition matrix of the time series of all GM voxels outside of the ROI). Next, a similarity matrix of functional connectivity within the ROI is computed and non-linear manifold learning using the Laplacian Eigenmaps algorithm is employed. The derived connectopic maps represent how connectivity between the FFG and the rest of the cortex varies topographically within the ROI. Furthermore, we also estimated a reference connectopic map using 20 unrelated individuals from the Human Connectome Project (HCP)^33^, which has excellent data quality. We correlated this reference connectopy with those resulting from LEAP to assess the quality of the obtained connectopies, as well as of the validity of the main axis of connectivity change (see also ^34^). We made sure that all included FFG gradients had a correlation of at least *r*=0.5 with the average reference gradient computed in the HCP sample.

### Sensitivity Analyses

#### Head Motion

To ensure that results were not confounded by head motion, we removed autistic male participants with the highest meanFD values until group differences in meanFD across diagnosis and sex were no longer significant (*p*>0.1). This resulted in a reduced sample of 497 individuals after excluding 17 autistic males (meanFD: χ²=6.15, *p*=0.1). All analyses were then repeated on this motion-balanced sample. Results remained significant for normalised counts (β=0.003, SE=0.001, 95% CI [0.0008, 0.005], *t*=2.66, *p*=0.008, *p*FDR=0.05), dwell time (β=0.025, SE=0.01, 95% CI [0.003, 0.046], *t*=2.25, *p*=0.025) and transition probabilities from baseline to μCAP3 (odds ratio [OR]=1.27, 95% CI [1.08, 1.49], *Z*=2.85, *p*=0.004, *p*FDR=0.04) and transition probabilities from μCAP3 to μCAP3 (OR=1.49, 95% CI [1.12, 1.99], *Z*=2.75, *p*=0.006, *p*FDR=0.04).

When reanalysed in the motion-balanced subsample, the sex-by-diagnosis interaction on μCAP3 spatial extent at the 90th percentile threshold remained trend-level (β=−0.001, SE<0.001, 95% CI [-0.003, 0.0001], *t*=−1.81, *p*=0.07), consistent in direction with the main analysis.

#### FIQ

Given differences in intellectual functioning across the groups, we reran analyses including FIQ as an additional covariate. When additionally controlling for FIQ, results remained significant for normalised counts (β=0.003, SE=0.001, 95% CI [0.001, 0.005], *t*=2.9, *p*=0.004, *pFDR*=0.023), dwell time (β=0.027, SE=0.001, 95% CI [0.005, 0.048], *t*=2.42, *p*=0.016) and transition probabilities from baseline to μCAP3 (OR=1.27, 95% CI [1.08, 1.5], *Z*=2.88, *p*=0.004, *p*FDR=0.03) and transition probabilities from μCAP3 to μCAP3 (OR=1.51, 95% CI [1.13, 2.01], *Z*=2.84, *p*=0.005, *p*FDR=0.03).

The sex-by-diagnosis interaction on the spatial extent of μCAP3 using a 90th percentile threshold to define suprathreshold voxels remained significant, too (β=−0.001, SE=0.001, 95% CI [-0.003, -0.0003], *t*=−2.01, *p*=0.045) and comparable across different thresholds.

### Supplementary Figures

#### Figure S1

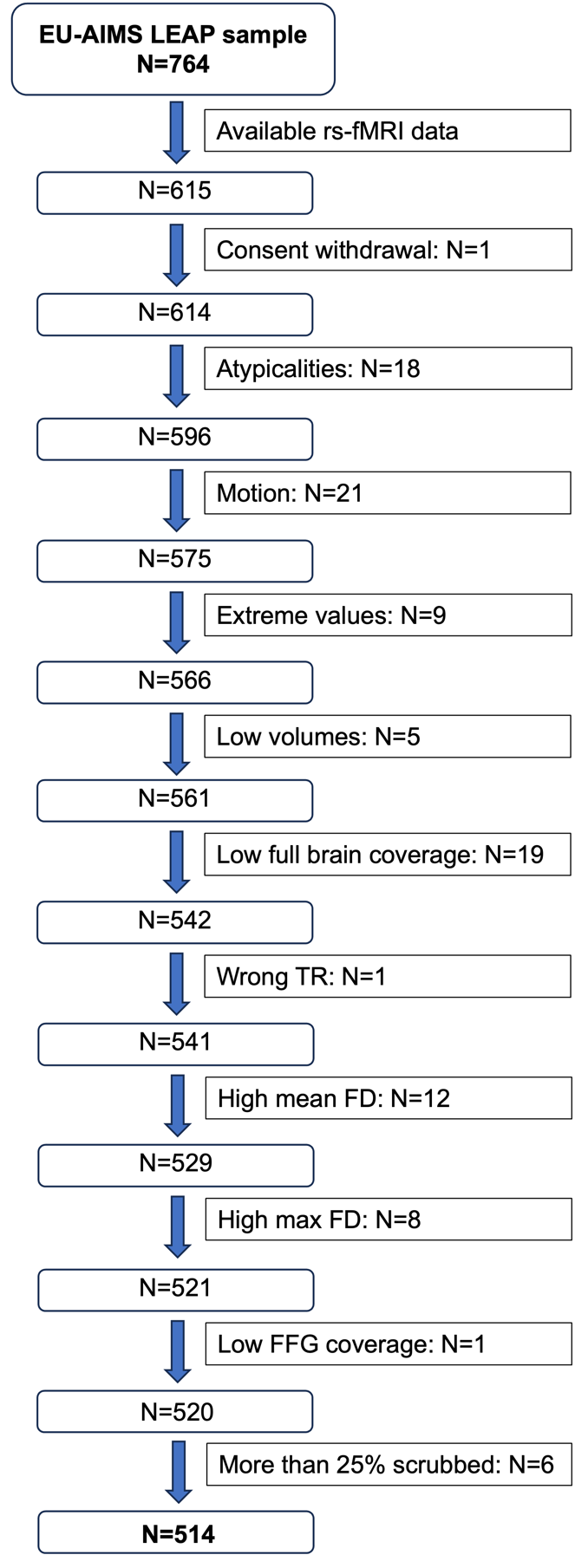

**Figure S1. Flowchart and** **exclusion criteria**. **Abbreviations**: rs-fMRI=resting-state fMRI; FD=framewise displacement in mm; FFG=fusiform gyrus.

#### Figure S2

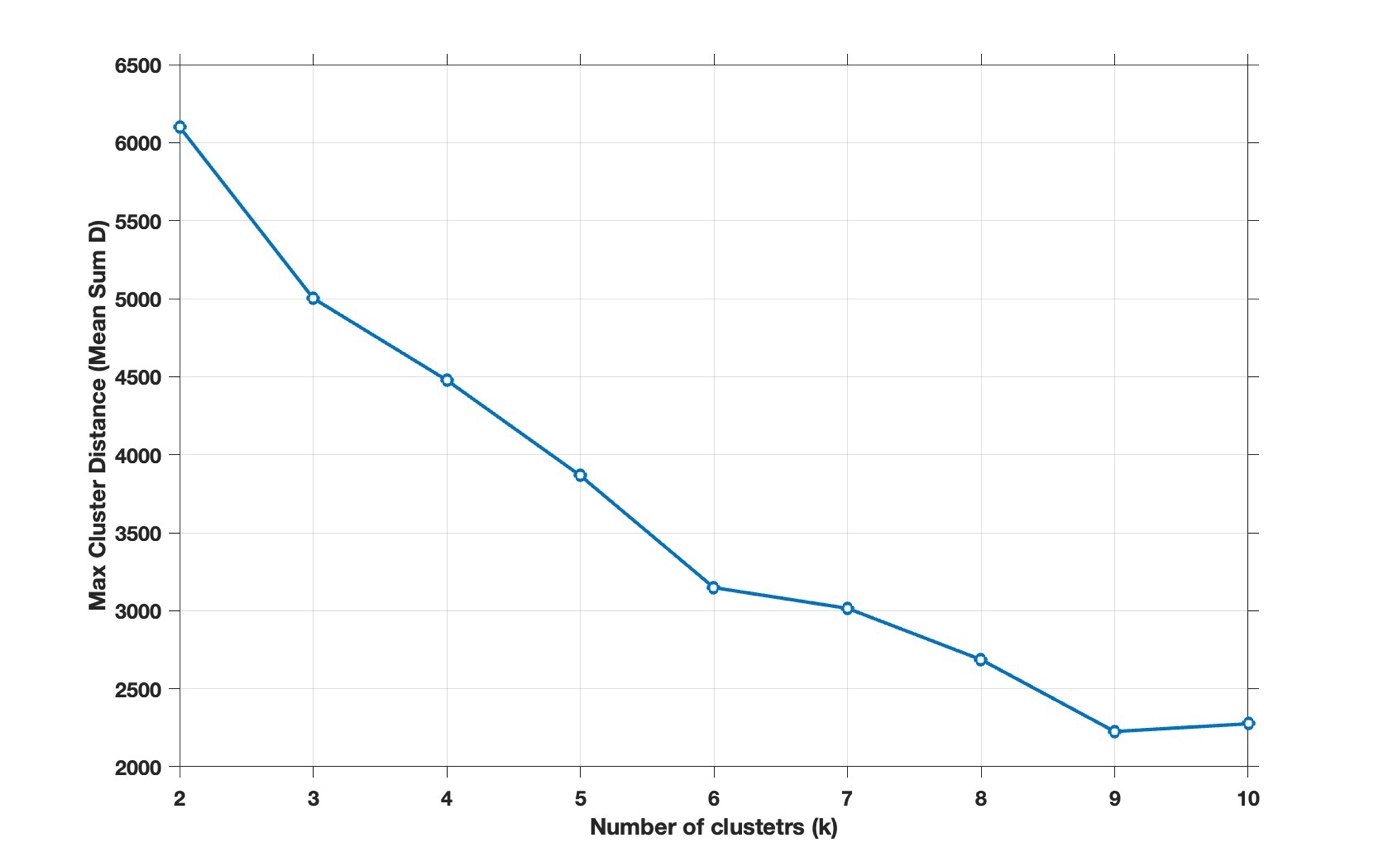

**Figure S2.** Sum of distances of test frames for the worst-fit cluster for each candidate K value within the range of 2 to 10. K=6 was chosen as the optimal value, as detailed in the Methods.

#### Figure S3

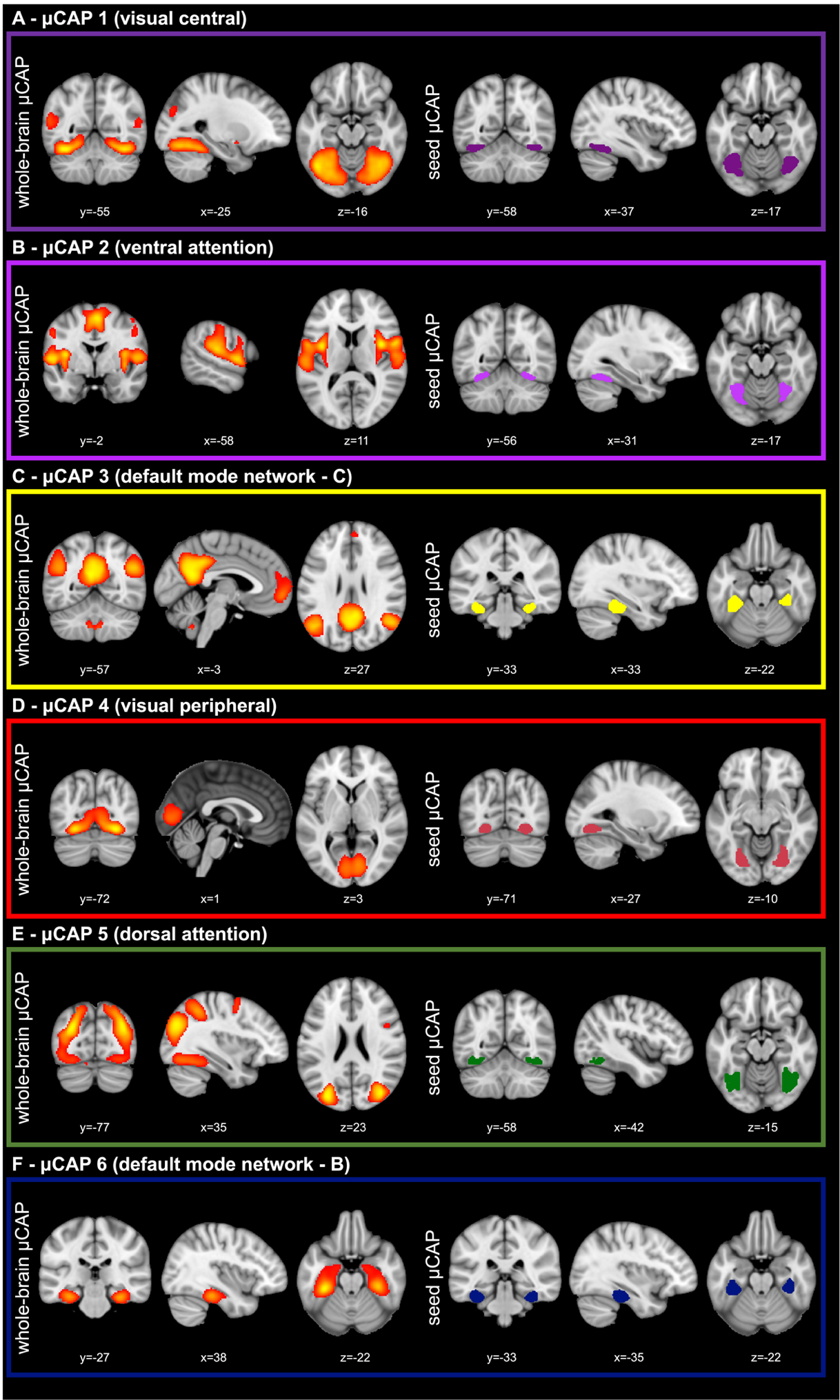

**Figure S3.** Each of the identified large-scale brain networks was associated with a specific data-driven subregion of FFG.  The left side of panels A–F show the spatial maps of the six μCAPs (thresholded at the 90th percentile):  a visual-central μCAP (A), a ventral attention μCAP (B), a default mode network (DMN) - C μCAP (C), a visual-peripheral μCAP (D), a dorsal attention μCAP (E) and a DMN - B μCAP (F). The right side of panels A–F shows the corresponding subregion of the FFG functionally mapping onto the whole-brain μCAP.

#### Figure S4

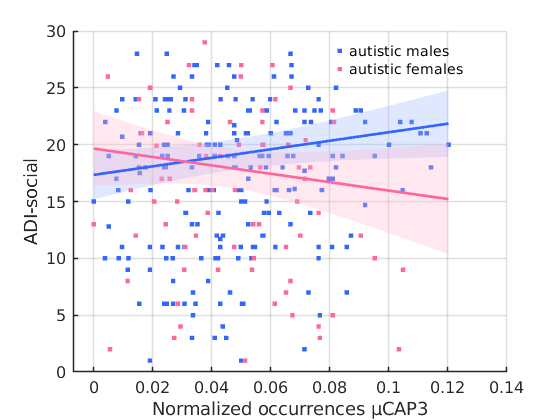

**Figure S4.** Association between normalised counts of μCAP3 and social symptoms as measured by the ADI-R in autistic individuals. Each point represents one participant (autistic males = blue; autistic females = pink).

#### Figure S5

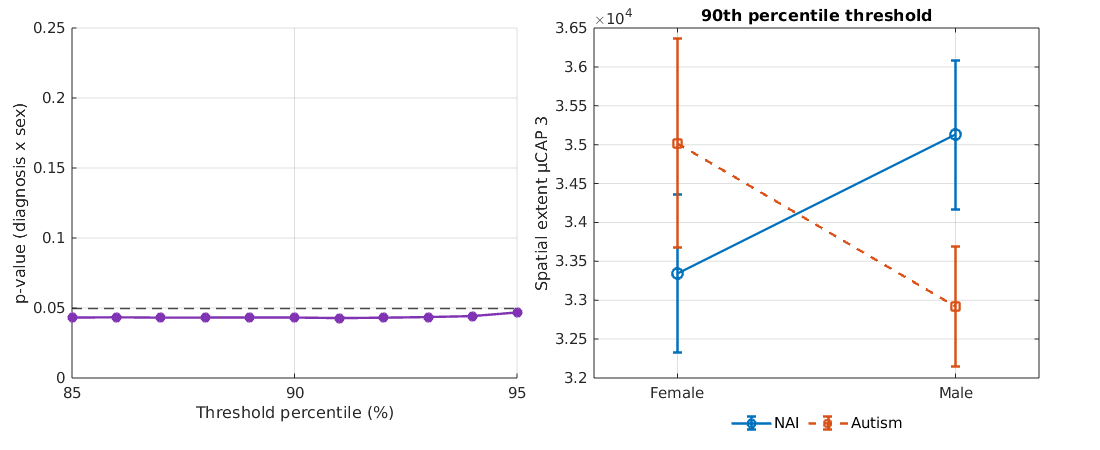

**Figure S5.** (A) *p*-values for the sex-by-diagnosis interaction on voxel-wise spatial extent within μCAP3, computed across multiple percentile thresholds (0.85–0.95, step=0.01). The dashed horizontal line marks the conventional *p* = 0.05 significance level. (B) Sex-by-diagnosis interaction at the 90th percentile threshold, showing group means and standard errors for spatial extent. Lines indicate opposite patterns of effect across sexes in autistic (orange, dashed) and non-autistic (blue, solid) participants.

### Supplementary Tables

#### Table S1

| Site | Ethics committee | ID/reference no. |
| --- | --- | --- |
| KCL, UCAM | London Queen Square Health Research Authority Research Ethics Committee | 13/LO/1156 |
| RUNMC, UMCU | Radboud Universitair Medisch Centrum | 2013/455 |
|  | Instituut Waarborging Kwaliteit en Veiligheid |  |
|  | Commissie Mensgebonden Onderzoek |  |
|  | Regio Arnhem-Nijmegen (Radboud University Medical Centre Institute Ensuring Quality and Safety |  |
|  | Committee on Research Involving Human Subjects |  |
|  | Arnhem-Nijmegen) |  |
| CIMH | UMM Universitätsmedizin Mannheim, Medizinische Ethik Commission II (UMM University Medical Mannheim, Medical Ethics Commission II) | 2014-540N-MA |

**Table S1.** Ethics approval of the included sites.

#### Table S2

| **Site** | **Manufacturer** | **Model** | **Software Version** | **Acquisition sequence** | **Coverage** | **Slices** | **Thickness [mm]** | **Resolution [mm^3^]** | **TR [s]** | **TE [ms]** | **FA [°]** | **FOV** |
| --- | --- | --- | --- | --- | --- | --- | --- | --- | --- | --- | --- | --- |
| Cambridge | Siemens | Verio | Syngo MR B17 | Tfl3d1_ns | 256*256 | 176 | 1.2 | 1.1*1.1*1.2 | 2.3 | 2.95 | 9 | 270 |
| London | GE Medical systems | Discovery mr750 | LX MR DV23.1_V02_1317.c | SAG ADNI GO ACC SPGR | 256*256 | 196 | 1.2 | 1.1*1.1*1.2 | 7.31 | 3.02 | 11 | 270 |
| Mannheim | Siemens | TimTrio | Syngo MR B17 | MPRAGE ADNI | 256*256 | 176 | 1.2 | 1.1*1.1*1.2 | 2.3 | 2.93 | 9 | 270 |
| Nijmegen | Siemens | Skyra | Syngo MRD13 | Tfl3d1_16ns | 256*256 | 176 | 1.2 | 1.1*1.1*1.2 | 2.3 | 2.93 | 9 | 270 |
| Rome | GE Medical systems | Signa HDxt | 24/LX/MR HD16.0_V02_1131.a | SAG ADNI GO ACC SPGR | 256*256 | 172 | 1.2 | 1.1*1.1*1.2 | 5.96 | 1.76 | 11 | 270 |
| Utrecht | Philips Medical Systems | Achieva/ | 3.2.3, 3.2.3.1 | ADNI GO 2 | 256*256 | 170 | 1.2 | 1.1*1.1*1.2 | 6.76 | 3.1 | 9 | 270 |
|  |  | Ingenia CX |  |  |  |  |  |  |  |  |  |  |

**Table S2.** Scanner-related parameters (structure).

#### Table S3

| **Site** | **Scanner** | **Field strength** | **Instruct.** | **TR /TE1/TE2 / TE3 (ms)/FA (°)** | **No. of vol.** | **No. of slices** | **Voxel size** |
| --- | --- | --- | --- | --- | --- | --- | --- |
| Cambridge | Siemens Magnetom Verio | 3T | Fixation | 2300/ 12/29/46/80 | 266 | 33 | 3.8 x 3.8 x 3.8 |
| KCL | GE NA | 3T | Fixation | 2300/ NA/31/48/90 | 215 | 33 | 3.8 x 3.8 x 3.8 |
| Mannheim | Siemens Magnetom TIM Trio | 3T | Fixation | 2300/ 12/29/46/80 | 215 | 33 | 3.8 x 3.8 x 3.8 |
| Nijmegen | Siemens Magnetom Skyra | 3T | Fixation | 2300/ 12/NA/NA/80 | 266 | 33 | 3.8 x 3.8 x 3.8 |
| Utrecht | Philips | 3T | Fixation | 2300/13/31/49/80 | 200 | 33 | 3.75 x 3.75 x 3.75 |

**Table S3.** Scanner-related parameters (function).

#### Table S4

| **Diagnostic group** | **Unknown**  **% (n)** | **No**  **% (n)** | **Yes**  **% (n)** | **Medication and Categories** |
| --- | --- | --- | --- | --- |
| autistic males  (n=208) | 6% (12) | 54% (112) | 40% (84)  One: n=45  Two: n=22  Three: n=5  Other medication use: n=12 | Antidepressants (n=17)   - Selective serotonin reuptake inhibitor (SSRI) (n=13) - Tetracyclic antidepressant (TeCA) (n=2) - Tricyclic Antidepressant (TCA) (n=2)   Antiepileptics (n=4; no additional information)  Antimigraine preparations (n=2; no additional information)  Gastro-intestinal / genito-urinary disorder medication   - Hyoscine Butylbromide (n=1)   Anxiolytics (n=1; no additional information)  Other analgesics and antipyretics (n=3)  Antipsychotics (n=19)   - Aripiprazole (n=3) - Risperidone (n=14) - Quetiapine (n=1) - Pipamperone (n=1)   Hypnotics and sedatives   - Melatonin (n=22)   Psychostimulants and other drugs used to treat ADHD (n=35)   - Atomoxetine (n=3) - Dexamfetamine (n=1) - Methylphenidate hydrochloride (n=31) |
| autistic females  (n=78) | 8% (6) | 46% (36) | 46% (36)  One: n=21  Two: n=4  Three: n=2  Other medication use: n=9 | Antidepressants (n=13)   - Selective serotonin reuptake inhibitor (SSRI) (n=12) - Tricyclic Antidepressant (TCA) (n=1)   Antiepileptics (n=3; no additional information)  Antimigraine preparations (n=1; no additional information)  Anxiolytics (n=1; no additional information)  Antipsychotics (n=4)   - Aripiprazole (n=2) - Risperidone (n=1) - Quetiapine (n=1)   Hypnotics and sedatives   - Melatonin (n=6)   Psychostimulants and other drugs used to treat ADHD   - Methylphenidate hydrochloride (n=7) |
| Non-autistic males  (n=146) | 8% (12) | 77% (113) | 14% (21)  One: n=11  Other medication use: n=10 | Antidepressants   - Selective serotonin reuptake inhibitor (SSRI) (n=1)   Antiepileptics (n=1; no additional information)  Other analgesics and antipyretics (n=3)  Hypnotics and sedatives   - Melatonin (n=1)   Psychostimulants and other drugs used to treat ADHD   - Methylphenidate hydrochloride (N=5) |
| Non-autistic females  (n=82) | 9% (7) | 61% (50) | 30% (25)  One: n=4  Two: n=1  Three: n=1  Other medication use: n=19 | Antidepressants   - Selective serotonin reuptake inhibitor (SSRI) (n=3) - Tetracyclic antidepressant (TeCA) (n=1)   Antipsychotics   - Risperidone (n=1)   Other analgesics and antipyretics (n=1)  Antiepileptics (n=1; no additional information)  Drugs used in addictive disorder (n=1; no additional information)  Psychostimulants and other drugs used to treat ADHD  Methylphenidate hydrochloride (N=1) |

**Table S4.** Information on medication use in the sample. Abbreviations: ADHD=attention-deficit/hyperactivity disorder

#### Table S5

| **CAP** | **Anatomical Region** | **Overlap Percentage** |
| --- | --- | --- |
| **μCAP1** | Temporal Occipital Fusiform Cortex | 86.52 |
|  | Occipital Fusiform Gyrus | 80.18 |
|  | Lingual Gyrus | 67.91 |
|  | Planum Temporale | 58.72 |
|  | Intracalcarine Cortex | 54.18 |
|  | Supracalcarine Cortex | 49.81 |
|  | Cuneal Cortex | 37.73 |
|  | Juxtapositional Lobule Cortex (formerly Supplementary Motor Cortex) | 29.11 |
|  | Lateral Occipital Cortex, inferior division | 27.93 |
|  | Central Opercular Cortex | 26.03 |
|  | Inferior Temporal Gyrus, temporooccipital part | 21.09 |
|  | Cingulate Gyrus, posterior division | 20.52 |
|  | Heschl's Gyrus (includes H1 and H2) | 20.36 |
|  | Parietal Operculum Cortex | 19.03 |
|  | Cingulate Gyrus, anterior division | 18.6 |
|  | Angular Gyrus | 15.69 |
|  | Middle Temporal Gyrus, temporooccipital part | 14.42 |
|  | Precuneous Cortex | 13.96 |
|  | Planum Polare | 13.35 |
|  | Precentral Gyrus | 11.58 |
|  | Temporal Fusiform Cortex, posterior division | 10.92 |
|  | Supramarginal Gyrus, posterior division | 10.71 |
|  | Superior Temporal Gyrus, posterior division | 8.2 |
|  | Insular Cortex | 5.98 |
|  | Postcentral Gyrus | 5.16 |
|  | Lateral Occipital Cortex, superior division | 5.09 |
|  | Superior Temporal Gyrus, anterior division | 3.26 |
|  | Temporal Pole | 2.89 |
|  | Occipital Pole | 2.86 |
|  | Paracingulate Gyrus | 2.75 |
|  | Inferior Frontal Gyrus, pars opercularis | 2.6 |
|  | Middle Frontal Gyrus | 1.67 |
|  | Parahippocampal Gyrus, posterior division | 1.13 |
|  | Supramarginal Gyrus, anterior division | 0.63 |
|  | Inferior Temporal Gyrus, posterior division | 0.1 |
| **μCAP2** | Central Opercular Cortex | 78.74 |
|  | Juxtapositional Lobule Cortex (formerly Supplementary Motor Cortex) | 75.61 |
|  | Parietal Operculum Cortex | 65.95 |
|  | Insular Cortex | 45.54 |
|  | Supramarginal Gyrus, anterior division | 44.78 |
|  | Planum Temporale | 39.71 |
|  | Temporal Occipital Fusiform Cortex | 33.83 |
|  | Frontal Operculum Cortex | 29.55 |
|  | Precentral Gyrus | 21.29 |
|  | Planum Polare | 18.56 |
|  | Postcentral Gyrus | 18.04 |
|  | Cingulate Gyrus, anterior division | 14.51 |
|  | Heschl's Gyrus (includes H1 and H2) | 12.42 |
|  | Inferior Frontal Gyrus, pars opercularis | 11.13 |
|  | Occipital Fusiform Gyrus | 9.86 |
|  | Paracingulate Gyrus | 5.1 |
|  | Supramarginal Gyrus, posterior division | 4.56 |
|  | Lingual Gyrus | 3.05 |
|  | Superior Frontal Gyrus | 2.95 |
|  | Temporal Pole | 2.53 |
|  | Superior Temporal Gyrus, anterior division | 2.3 |
|  | Cingulate Gyrus, posterior division | 2.11 |
|  | Temporal Fusiform Cortex, posterior division | 1.76 |
|  | Superior Temporal Gyrus, posterior division | 1.31 |
|  | Superior Parietal Lobule | 0.96 |
|  | Lateral Occipital Cortex, inferior division | 0.51 |
|  | Middle Temporal Gyrus, temporooccipital part | 0.45 |
|  | Middle Frontal Gyrus | 0.04 |
|  | Precuneous Cortex | 0.02 |
| **μCAP3** | Cingulate Gyrus, posterior division | 49.05 |
|  | Parahippocampal Gyrus, posterior division | 45.92 |
|  | Precuneous Cortex | 39.93 |
|  | Angular Gyrus | 32.38 |
|  | Temporal Fusiform Cortex, posterior division | 27.98 |
|  | Middle Temporal Gyrus, anterior division | 26.18 |
|  | Lateral Occipital Cortex, superior division | 23.55 |
|  | Temporal Occipital Fusiform Cortex | 16 |
|  | Paracingulate Gyrus | 14.23 |
|  | Superior Frontal Gyrus | 11.2 |
|  | Frontal Medial Cortex | 10.66 |
|  | Middle Temporal Gyrus, posterior division | 6.81 |
|  | Middle Frontal Gyrus | 6.09 |
|  | Frontal Pole | 5.83 |
|  | Superior Temporal Gyrus, anterior division | 4.41 |
|  | Supracalcarine Cortex | 2.3 |
|  | Lingual Gyrus | 2.13 |
|  | Cuneal Cortex | 1.71 |
|  | Intracalcarine Cortex | 1.57 |
|  | Superior Temporal Gyrus, posterior division | 1.2 |
|  | Parahippocampal Gyrus, anterior division | 1.04 |
|  | Middle Temporal Gyrus, temporooccipital part | 0.9 |
|  | Supramarginal Gyrus, posterior division | 0.49 |
| **μCAP4** | Intracalcarine Cortex | 60.74 |
|  | Occipital Fusiform Gyrus | 52.44 |
|  | Supracalcarine Cortex | 49.81 |
|  | Lingual Gyrus | 43.66 |
|  | Temporal Occipital Fusiform Cortex | 23.76 |
|  | Cuneal Cortex | 10.68 |
|  | Occipital Pole | 8.02 |
|  | Lateral Occipital Cortex, superior division | 5.42 |
|  | Lateral Occipital Cortex, inferior division | 0.27 |
| **μCAP5** | Temporal Occipital Fusiform Cortex | 69.64 |
|  | Occipital Fusiform Gyrus | 55.2 |
|  | Lateral Occipital Cortex, superior division | 46.96 |
|  | Inferior Temporal Gyrus, temporooccipital part | 36.94 |
|  | Lateral Occipital Cortex, inferior division | 36.73 |
|  | Superior Parietal Lobule | 34.8 |
|  | Middle Temporal Gyrus, temporooccipital part | 7.09 |
|  | Temporal Fusiform Cortex, posterior division | 6.21 |
|  | Lingual Gyrus | 4.97 |
|  | Supramarginal Gyrus, posterior division | 4.96 |
|  | Middle Frontal Gyrus | 4.04 |
|  | Supramarginal Gyrus, anterior division | 3.56 |
|  | Occipital Pole | 3.13 |
|  | Inferior Frontal Gyrus, pars opercularis | 2.38 |
|  | Precuneous Cortex | 1.83 |
|  | Precentral Gyrus | 1.71 |
|  | Superior Frontal Gyrus | 1.65 |
|  | Cuneal Cortex | 1.47 |
|  | Parahippocampal Gyrus, posterior division | 1.13 |
|  | Angular Gyrus | 0.42 |
|  | Postcentral Gyrus | 0.35 |
| **μCAP6** | Temporal Fusiform Cortex, posterior division | 55.83 |
|  | Parahippocampal Gyrus, posterior division | 42.39 |
|  | Parahippocampal Gyrus, anterior division | 12.74 |
|  | Temporal Occipital Fusiform Cortex | 10.28 |
|  | Inferior Temporal Gyrus, posterior division | 4.84 |
|  | Inferior Temporal Gyrus, temporooccipital part | 0.41 |
|  | Lingual Gyrus | 0.15 |

**Table S5.** Overlap of each μCAP with subregions of the Harvard-Oxford template.

#### Table S6

|  | **Sex Differences in NAI** | | **Sex Differences in ASC** | |
| --- | --- | --- | --- | --- |
| **Bin** | **p** | **q** | **p** | **q** |
| **Right FFG** |  |  |  |  |
| 1 | 0.014642 | 0.23029 | 0.18779 | 0.36777 |
| 2 | 0.025251 | 0.23029 | 0.22719 | 0.37865 |
| 3 | 0.057571 | 0.23029 | 0.30523 | 0.43605 |
| 4 | 0.18102 | 0.36777 | 0.46994 | 0.58743 |
| 5 | 0.62049 | 0.72999 | 0.78901 | 0.78901 |
| 6 | 0.62049 | 0.72999 | 0.78901 | 0.78901 |
| 7 | 0.18102 | 0.36777 | 0.46994 | 0.58743 |
| 8 | 0.057571 | 0.23029 | 0.30523 | 0.43605 |
| 9 | 0.025251 | 0.23029 | 0.22719 | 0.37865 |
| 10 | 0.014642 | 0.23029 | 0.18779 | 0.36777 |
| **Left FFG** |  |  |  |  |
| 1 | 1.91E-06 | 3.81E-05 | 0.43735 | 0.62479 |
| 2 | 1.26E-05 | 0.00012635 | 0.47641 | 0.63521 |
| 3 | 0.00021048 | 0.0014032 | 0.54546 | 0.68182 |
| 4 | 0.009027 | 0.045135 | 0.67004 | 0.74449 |
| 5 | 0.33371 | 0.55618 | 0.87462 | 0.87462 |
| 6 | 0.33371 | 0.55618 | 0.87462 | 0.87462 |
| 7 | 0.009027 | 0.045135 | 0.67004 | 0.74449 |
| 8 | 0.00021048 | 0.0014032 | 0.54546 | 0.68182 |
| 9 | 1.26E-05 | 0.00012635 | 0.47641 | 0.63521 |
| 10 | 1.91E-06 | 3.81E-05 | 0.43735 | 0.62479 |

**Table S6.** Abbreviations: NAI=non-autistic individuals, ASC=autism spectrum condition

#### Table S7

**Table S7.** Statistics for effects of diagnosis, sex and diagnosis-by-sex interaction for normalised occurrences.

|  | **Diagnosis** | | | | | | | **Sex** | | | | | | | **Diagnosis-by-Sex** | | | | | | |
| --- | --- | --- | --- | --- | --- | --- | --- | --- | --- | --- | --- | --- | --- | --- | --- | --- | --- | --- | --- | --- | --- |
|  | beta | CI low | CI high | SE | t | p | pFDR | beta | CI low | CI high | SE | t | p | pFDR | beta | CI low | CI high | SE | t | p | pFDR |
| μCAP1 | -0.0002 | -0.0046 | 0.0041 | 0.002 | -0.108 | 0.914 | 0.925 | 0.0071 | 0.0028 | 0.0113 | 0.002 | 3.270 | 0.001 | 0.007 | 0.0002 | -0.0041 | 0.0045 | 0.002 | 0.088 | 0.930 | 0.992 |
| μCAP2 | -0.0012 | -0.0037 | 0.0012 | 0.001 | -1.006 | 0.315 | 0.630 | -0.0028 | -0.0051 | -0.0004 | 0.001 | -2.302 | 0.022 | 0.041 | 0.0013 | -0.0011 | 0.0037 | 0.001 | 1.075 | 0.283 | 0.849 |
| μCAP3 | 0.0005 | -0.0017 | 0.0027 | 0.001 | 0.423 | 0.672 | 0.925 | -0.0028 | -0.0050 | -0.0006 | 0.001 | -2.466 | 0.014 | 0.041 | **0.0032** | **0.0010** | **0.0054** | **0.001** | **2.88** | **0.004** | **0.025** |
| μCAP4 | 0.0017 | -0.0007 | 0.0040 | 0.001 | 1.370 | 0.171 | 0.607 | -0.0026 | -0.0050 | -0.0003 | 0.001 | -2.211 | 0.027 | 0.041 | -0.0004 | -0.0027 | 0.0020 | 0.001 | -0.30 | 0.766 | 0.992 |
| μCAP5 | -0.0001 | -0.0025 | 0.0023 | 0.001 | -0.094 | 0.925 | 0.925 | -0.0017 | -0.0041 | 0.0007 | 0.001 | -1.421 | 0.156 | 0.187 | 1.20E-05 | -0.0024 | 0.0024 | 0.001 | 0.010 | 0.992 | 0.992 |
| μCAP6 | 0.0015 | -0.0008 | 0.0038 | 0.001 | 1.276 | 0.202 | 0.607 | 0.0001 | -0.0021 | 0.0024 | 0.001 | 0.101 | 0.920 | 0.920 | 0.0003 | -0.0020 | 0.0025 | 0.001 | 0.233 | 0.816 | 0.992 |
